# Causation Between Smoking Quantity and Depressive Symptoms in Young Adults: Evidence From Novel Cross-Lagged Twin Models

**DOI:** 10.1101/2025.11.18.25340516

**Authors:** Madhurbain Singh, Michael D. Hunter, Elham Assary, Brad Verhulst, Roseann E. Peterson, Hermine H. M. Maes, Conor V. Dolan, Thalia C. Eley, Michael C. Neale

## Abstract

**Background:** Smoking and depression often co-occur in young adults, potentially due to reciprocal causal effects and/or shared underlying etiological factors. Here, we tested causal hypotheses between smoking quantity (cigarettes per day; *CigDay*) and depressive symptoms (*DepSx*) using novel Biometrical Cross-Lagged Panel Models (CLPMs), which integrate twin developmental and direction-of-causation analyses for more robust causal inference in longitudinal twin studies.

**Methods:** Study sample included 10,034 participants (61.6% females; 4,112 twin pairs) from the Twins Early Development Study, with up to six repeated assessments spanning ages 21–29. The Biometrical CLPM provided three key innovations over standard twin CLPM: distinct autoregressive processes of latent genetic and environmental factors; cross-lagged genetic and environmental liabilities, contrasting confounding and causation; and cross-sectional causal effects, distinguishing between proximal (short-term) and distal (lagged) causation.

**Results:** In the two largest waves assessed approximately four years apart, genetic liabilities for both variables remained highly stable, while environmental influences showed considerable temporal variation. In the standard CLPM, *CigDay* predicted *DepSx* four years later. However, in the Biometrical CLPM, the cross-lagged phenotypic associations were attenuated and non-significant when accounting for the differential genetic and environmental developmental processes. Adding the cross-sectional causal paths revealed significant bidirectional *proximal* effects, with a stronger *CigDay→DepSx* effect. Across all six waves with intervals up to two years, analyses consistently showed significant bidirectional cross-lagged associations.

**Conclusions:** The findings support a reinforcing feedback loop between *CigDay* and *DepSx* during young adulthood, involving bidirectional effects that persist for up to two years but dissipate over longer intervals.

## Introduction

Young adulthood is a critical period for the development of psychopathology and substance use, with cigarette smoking and depressive symptoms increasing in prevalence (Kessler et al., 2005) and often co-occurring (Conway et al., 2018; Mathew et al., 2017). In population-based studies, higher depressive symptoms are associated with an increased risk of smoking (Weinberger et al., 2017). Likewise, smoking predicts subsequent depressive symptoms, suggesting reciprocal causal effects (Fluharty et al., 2016). However, this co-occurrence may also arise from correlated underlying familial (genetic and environmental) and individual-specific environmental influences (Lyons et al., 2008; McCaffery et al., 2008; Ranjit, Korhonen, et al., 2019). Identifying the mechanisms underlying the association between smoking and depressive symptoms is essential for evidence-informed preventative efforts.

Tests of causal hypotheses between smoking and depression using observational study designs have provided stronger support for the effects of smoking on depression than *vice versa*. For example, a longitudinal study of young adults using cross-lagged panel models (CLPMs) indicated that smoking predicts depression two years later, but not *vice versa*, suggesting a unidirectional causal process (Lee et al., 2023). Causal hypotheses have also been tested in *cross-sectional* data from monozygotic (MZ) and dizygotic (DZ) twins, leveraging cross-twin cross-trait correlations in the Direction-of-Causation (DoC) models (Heath et al., 1993). For instance, in a study of adult male twins, the DoC model with causal effects of nicotine dependence on major depressive disorder (MDD) fit the data slightly better than the model with only correlated genetic liabilities between the two (Edwards & Kendler, 2012).

Previous *longitudinal* twin studies did not support a causal relationship between depression and smoking status in emerging adulthood, as the prospective associations were primarily explained by familial confounding (Ranjit, Buchwald, et al., 2019; Ranjit, Korhonen, et al., 2019). However, these studies were restricted to discordant twin pairs, limiting the sample size and statistical power. Moreover, results based on separate regression models for each causal direction are likely to be biased if the true causal process is bidirectional. Here, we aimed to analyze the relationship between smoking and depression using CLPMs applied to twin data (Burt et al., 2005; Morneau-Vaillancourt et al., 2023; Van Bergen et al., 2025). Our combination of the twin CLPM with the DoC model confers several advantages outlined below.

First, the standard twin CLPM (Morneau-Vaillancourt et al., 2023) models temporal stability as a phenotypic autoregressive process, assuming that the *manifest phenotype* mediates the construct stability (i.e., autocorrelation) over time. In contrast, the *genetic simplex* model, which we incorporate in our model, decomposes the phenotypic processes into distinct genetic and environmental autoregressive processes (Boomsma & Molenaar, 1987). An application of this model to internalizing symptoms across adulthood revealed markedly different levels of stability in the genetic and environmental influences, with mean autocorrelations of 0.92 and 0.60, respectively, across 2-year age-bins (Nivard et al., 2015). This difference in genetic and environmental autocorrelations has implications for causal inference, as we demonstrate below. We incorporate the *genetic simplex* into our twin CLPM to model the stability and change in the genetic and environmental factors underlying each construct, and examine its impact on the causal inference based on prospective cross-lagged phenotypic associations.

Second, inferring causality from the cross-lagged associations in CLPM requires assuming that there is no cross-lagged confounding, which is untestable in data from unrelated individuals. However, in twin data, we can compare the standard twin CLPM (i.e., “causation” model) with a model including correlated cross-lagged genetic and environmental liabilities, similar to the cross-sectional twin causal analyses. Testing the alternative sources of cross-lagged associations in twin CLPM may provide more robust causal evidence if the “causation” model indeed yields the most parsimonious fit.

Finally, the prospective associations in CLPM typically decay with increasing intervals between assessments (Singh et al., 2024), becoming negligible at longer intervals. In one prior CLPM (Lee et al., 2023), the non-significant lagged effect of depression on smoking two years later may mask undetected short-term causation. To address this limitation, previous studies of unrelated individuals have integrated instrumental variables in CLPM to estimate both cross-sectional (“proximal” or short-term) and lagged (“distal”) effects (Singh et al., 2024). By combining the DoC twin model and the CLPM, we can estimate such cross-sectional effects without including instrumental variables.

We tested for bidirectional causal effects between smoking quantity and depressive symptoms in 10,034 young-adult participants (including 4,112 twin pairs) from the Twins Early Development Study (Lockhart et al., 2023; Rimfeld et al., 2019). To address the limitations of standard twin CLPM, we developed the novel *Biometrical CLPM,* incorporating three key innovations:

1. Allowing for differential developmental processes in the genetic and environmental factors underlying each construct;
2. Testing different hypotheses about the sources of cross-lagged associations: causation versus correlated genetic and environmental liabilities; and
3. Estimating proximal effects by integrating the principles of DoC model in CLPM, in addition to the distal effects.

The results indicate potential bidirectional short-term causal effects between smoking quantity and depressive symptoms and highlight the insights provided by the novel Biometrical CLPMs.

## Methods

The study was pre-registered on June 11, 2024 (https://doi.org/10.17605/OSF.IO/K47BF). Deviations from the preregistration are described in **Supplementary Note**.

### Study Sample

TEDS is an ongoing population-based prospective cohort study of twins born in England and Wales in 1994-96 (Lockhart et al., 2023). During young adulthood, over 8,000 twin pairs were invited to participate across six repeated assessments (in 2018-22), two of which were core study waves: “TEDS21” in 2018, N=8,005, aged 21-26 (Rimfeld et al., 2019), and “TEDS26” in 2021-22, N=7,927, aged 25-29 (Lockhart et al., 2023). After the start of the COVID-19 pandemic, four additional waves of data (“COVID Study, Phase 1–4”) were collected in April (N=4,596), July (N=3,800), October (N=3,470) 2020, and March 2021 (N=3,744) (Rimfeld et al., 2022). As the COVID Study waves have roughly half the sample size, we used data from TEDS21 and TEDS26 in our primary analyses, followed by analyses of all six waves to assess the consistency of results. The total sample comprised 10,034 participants with at least one assessment of smoking quantity (N=6,326) or depressive symptoms (N=10,003) across the six waves. The participants were 61.6% females and included 4,112 complete twin pairs: 1,042 MZ female, 531 MZ male, 903 DZ female, 426 DZ male, and 1,210 DZ opposite-sex pairs. The samples are described in **Supplementary Methods** and **Table 1**.

**Table 1.**
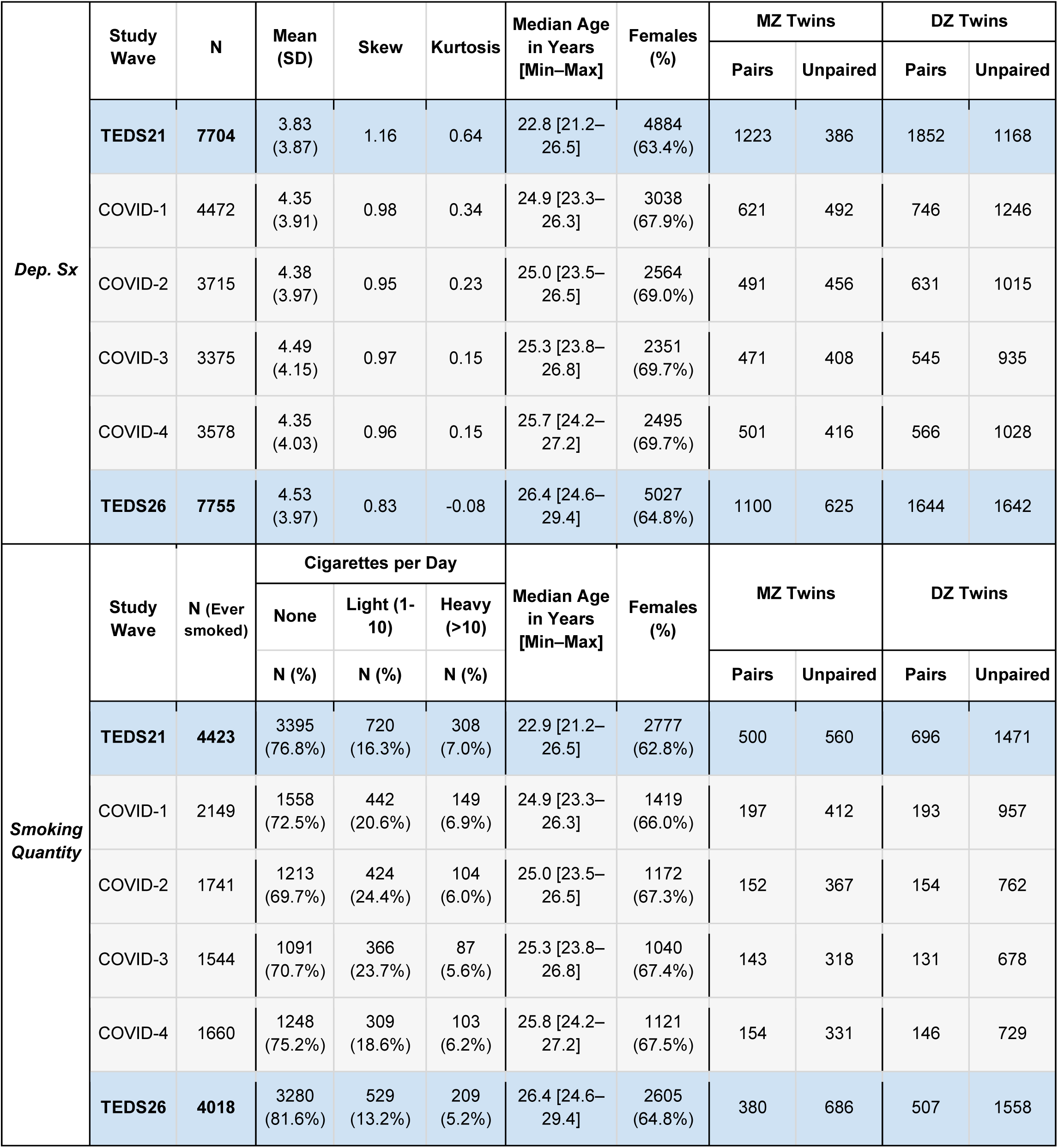
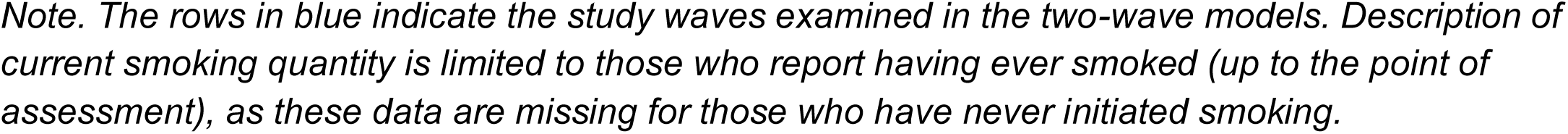
Sample Size and Descriptive Statistics by Measure and Study Wave.

|  | Study Wave | N | Mean (SD) | Skew | Kurtosis | Median Age in Years [Min–Max] | Females (%) | MZ Twins |  | DZ Twins |  |
| --- | --- | --- | --- | --- | --- | --- | --- | --- | --- | --- | --- |
|  |  |  |  |  |  |  |  | Pairs | Unpaired | Pairs | Unpaired |
| *Dep. Sx* | **TEDS21** | **7704** | 3.83 (3.87) | 1.16 | 0.64 | 22.8 [21.2–26.5] | 4884 (63.4%) | 1223 | 386 | 1852 | 1168 |
|  | COVID-1 | 4472 | 4.35 (3.91) | 0.98 | 0.34 | 24.9 [23.3–26.3] | 3038 (67.9%) | 621 | 492 | 746 | 1246 |
|  | COVID-2 | 3715 | 4.38 (3.97) | 0.95 | 0.23 | 25.0 [23.5–26.5] | 2564 (69.0%) | 491 | 456 | 631 | 1015 |
|  | COVID-3 | 3375 | 4.49 (4.15) | 0.97 | 0.15 | 25.3 [23.8–26.8] | 2351 (69.7%) | 471 | 408 | 545 | 935 |
|  | COVID-4 | 3578 | 4.35 (4.03) | 0.96 | 0.15 | 25.7 [24.2–27.2] | 2495 (69.7%) | 501 | 416 | 566 | 1028 |
|  | **TEDS26** | **7755** | 4.53 (3.97) | 0.83 | -0.08 | 26.4 [24.6–29.4] | 5027 (64.8%) | 1100 | 625 | 1644 | 1642 |
| *Smoking Quantity* | Study Wave | N (Ever smoked) | Cigarettes per Day |  |  | Median Age in Years [Min–Max] | Females (%) | MZ Twins |  | DZ Twins |  |
|  |  |  | None | Light (1-10) | Heavy (>10) |  |  |  |  |  |  |
|  |  |  | N (%) | N (%) | N (%) |  |  | Pairs | Unpaired | Pairs | Unpaired |
|  | **TEDS21** | **4423** | 3395 (76.8%) | 720 (16.3%) | 308 (7.0%) | 22.9 [21.2–26.5] | 2777 (62.8%) | 500 | 560 | 696 | 1471 |
|  | COVID-1 | 2149 | 1558 (72.5%) | 442 (20.6%) | 149 (6.9%) | 24.9 [23.3–26.3] | 1419 (66.0%) | 197 | 412 | 193 | 957 |
|  | COVID-2 | 1741 | 1213 (69.7%) | 424 (24.4%) | 104 (6.0%) | 25.0 [23.5–26.5] | 1172 (67.3%) | 152 | 367 | 154 | 762 |
|  | COVID-3 | 1544 | 1091 (70.7%) | 366 (23.7%) | 87 (5.6%) | 25.3 [23.8–26.8] | 1040 (67.4%) | 143 | 318 | 131 | 678 |
|  | COVID-4 | 1660 | 1248 (75.2%) | 309 (18.6%) | 103 (6.2%) | 25.8 [24.2–27.2] | 1121 (67.5%) | 154 | 331 | 146 | 729 |
|  | **TEDS26** | **4018** | 3280 (81.6%) | 529 (13.2%) | 209 (5.2%) | 26.4 [24.6–29.4] | 2605 (64.8%) | 380 | 686 | 507 | 1558 |

*Note. The rows in blue indicate the study waves examined in the two-wave models. Description of current smoking quantity is limited to those who report having ever smoked (up to the point of assessment), as these data are missing for those who have never initiated smoking.*

### Measures

#### Smoking Quantity: Cigarettes per Day (*CigDay*)

At each wave, participants were asked if they had ever smoked a cigarette, if they were currently smoking, and, if so, how many cigarettes they smoked per day (abbreviated as *CigDay*), yielding three levels of current smoking: *None*, *Light* (up to 10 cigarettes per day), and *Heavy* (more than 10 cigarettes per day; **Supplementary Methods**; **Table 1**). Since smoking initiation and quantity have distinct genetic and environmental liabilities (Bares et al., 2015; Do et al., 2015), *CigDay* had missing values for those who had never initiated smoking. There was no evidence for associations of *CigDay* with age and sex (**Supplementary Methods**). This ordinal variable was examined using the liability threshold model, assuming a normally distributed latent liability (Falconer, 1965).

#### Depressive Symptoms (*DepSx*)

In TEDS21 and COVID Study waves, depressive symptoms over the preceding two weeks were assessed using an 8-item abbreviated version of the 13-item *Short Mood and Feelings Questionnaire* (Angold et al., 1995). A sum score (range 0–16) was obtained if at least four (50%) items were answered. In TEDS26, the full 13-item sMFQ was assessed. However, for consistency, we computed the sum scores based on the eight items assessed in earlier waves. Total sMFQ scores were right-skewed (**Table 1**); therefore, these scores were regressed on age and sex, followed by a rank-based inverse-normal transformation of the residuals (**Supplementary Methods**). The transformed scores (*DepSx*) indicate an individual’s *liability* for depressive symptoms relative to their peers.

### Statistical Models

We initially fitted separate univariate biometrical (ACE) twin models to each assessment of *CigDay* and *DepSx* to estimate their additive genetic (A), familial environmental (C), and individual-specific environmental (E; including measurement error) variance components (Neale & Cardon, 1992) (**Supplementary Figure S1**).

#### Standard Twin CLPM

We fitted the standard two-wave twin CLPM to TEDS21 (wave 1) and TEDS26 (wave 2) data (**Figure 1A**), including cross-lagged paths between *DepSx* and *CigDay* (𝑏_𝑌_*_2_*_𝑋_*_1_* and 𝑏_𝑋_*_2_*_𝑌_*_1_*) and phenotypic autoregression (𝑎𝑟𝑋 and 𝑎𝑟𝑌). Similar to the univariate twin model, this model includes ACE variance decomposition of the phenotypic variances (e.g., 𝑉𝐴_𝑋_*_1_* , 𝑉𝐶_𝑋_*_1_*, and 𝑉𝐸_𝑋_*_1_* for *DepSx* at wave 1) and cross-sectional covariance at each wave (e.g., 𝑐𝑜𝑣𝐴*1*, 𝑐𝑜𝑣𝐶*1*, and 𝑐𝑜𝑣𝐸*1* between *DepSx* and *CigDay* at wave 1; **Supplementary Methods**).

**Figure 1.**
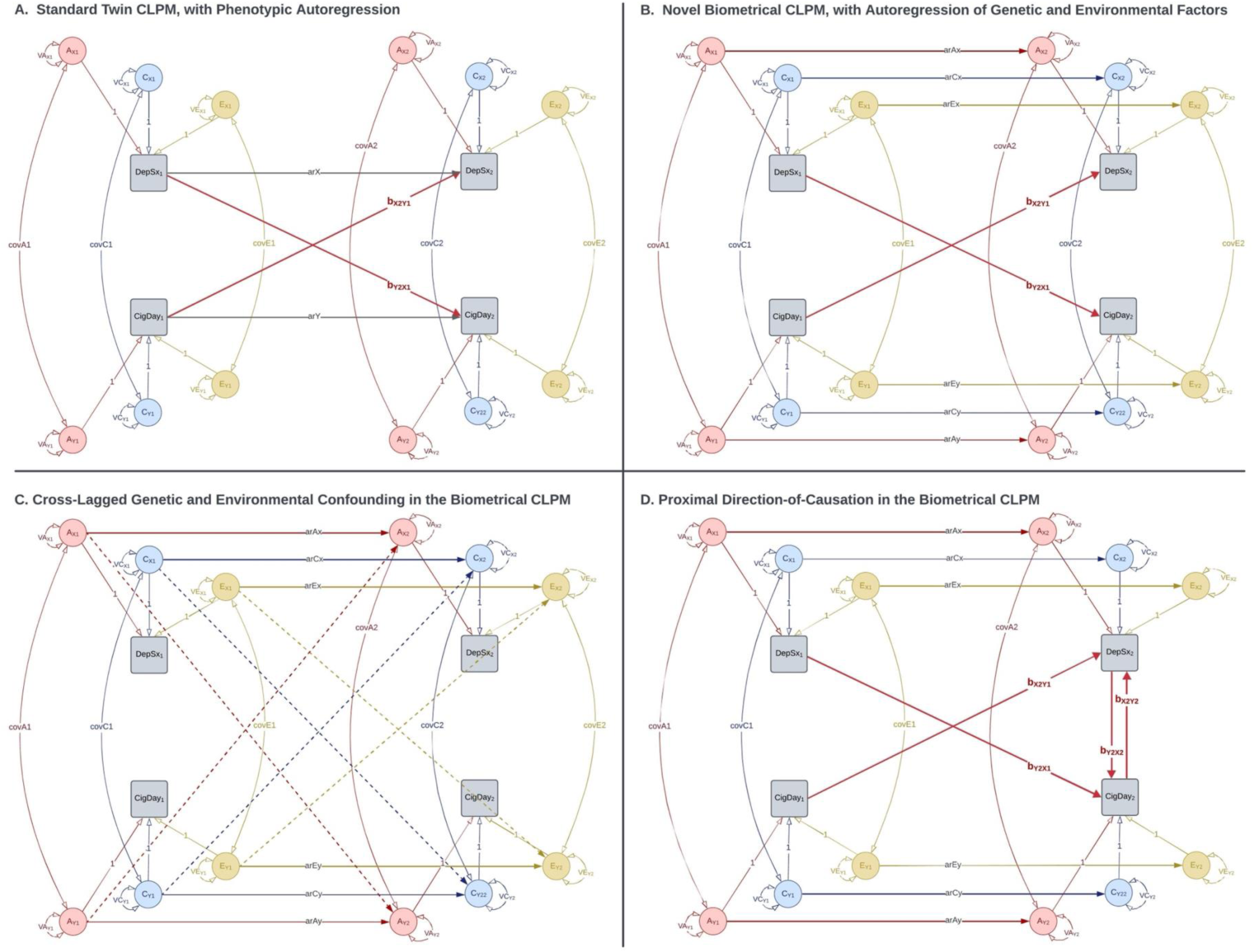
Cross-Lagged Twin Models. Note. These diagrams depict the cross-lagged model for one individual out of a twin pair. The variance of each observed phenotype is decomposed into latent additive genetic (A), shared familial environmental (C), and unique individual-specific environmental (E) factors. The twin correlation of the A factors is fixed at 1 in MZ and 0.5 in DZ twin pairs. The twin correlation of the C factors is fixed at 1 in both MZ and DZ pairs, while the twin correlation of the E factors is fixed at 0. The squares/rectangles indicate observed variables, the circles indicate latent variables, the single-headed arrows indicate regression paths, and the double-headed curved arrows indicate (co-)variances. Figure 1D shows only one of the different versions of this model with cross-sectional causal effects and/or confounding.

#### Biometrical Twin CLPM

In the novel Biometrical CLPM (**Figure 1B**), we replaced the phenotypic autoregressive paths with the freely-estimated ACE autoregressive paths; i.e., 𝑎𝑟𝐴𝑥, 𝑎𝑟𝐶𝑥, and 𝑎𝑟𝐸𝑥 between the respective latent factors of *DepSx* at waves 1 and 2, and 𝑎𝑟𝐴𝑦, 𝑎𝑟𝐶𝑦, and 𝑎𝑟𝐸𝑦 between the A, C, and E factors of *CigDay*. The standard twin CLPM is a nested, more restricted version of the Biometrical CLPM. If the standard model fits the data significantly worse than the Biometrical CLPM, it implies differential levels of autocorrelation of the A, C, and E (including measurement error) factors, which is inconsistent with stability as a phenotypic process. We examined the cross-lagged associations in the best-fitting model.

We then fitted an alternative model with cross-lagged confounding due to correlated genetic and environmental liabilities (**Figure 1C**). In this model, we replaced the phenotypic cross-lagged paths with cross-lagged paths between A, C, and E factors, e.g., a path from 𝐴_𝑋_*_1_* (*DepSx* at wave 1) to 𝐴_𝑌_*_2_* (*CigDay* at wave 2). Similar to the comparison of phenotypic and ACE autoregression, the model with phenotypic cross-lagged paths (the *causation* model) is nested within the model with cross-lagged ACE paths. The finding that the causation model fits as well as the cross-lagged ACE model supports rejecting the hypothesis of cross-lagged confounding and strengthens the evidence for cross-lagged causal effects.

Thus, using the Biometrical CLPM, we tested two alternative hypotheses concerning the cross-lagged associations: phenotypic causation (the hypothesis tested in the standard CLPM) and cross-lagged genetic and environmental confounding. These hypotheses were tested against each other and against the “null hypothesis” of no cross-lagged associations.

#### Biometrical CLPM-DoC: Proximal Direction-of-Causation

We examined cross-sectional proximal effects by applying the twin DoC model (Heath et al., 1993; Neale & Cardon, 1992) within the CLPM. If the two phenotypes differ in their ACE variance decomposition (i.e., different proportions of ACE variance components in the two phenotypes), DoC models with unidirectional or bidirectional phenotypic causal paths and models with only ACE covariance imply different levels of cross-twin cross-trait correlations. Comparing their goodness-of-fit statistics can help select the model that best fits the observed data. Hybrid DoC models combine phenotypic causation with correlated genetic and environmental liabilities (Maes et al., 2021). For instance, if both phenotypes have ACE variance decomposition, there are two potential phenotypic causal paths and three sources of confounding: 𝑐𝑜𝑣𝐴, 𝑐𝑜𝑣𝐶, and 𝑐𝑜𝑣𝐸. In a hybrid DoC model, we may estimate up to three of the five sources of covariance. If both phenotypes have AE variance decomposition, we may fit hybrid DoC models with any two sources of covariance, e.g., unidirectional causation and 𝑐𝑜𝑣𝐴.

We applied cross-sectional DoC and hybrid DoC models at wave 2 within the Biometrical CLPM, e.g., a model with bidirectional proximal causal effects (𝑏_𝑌_*_2_*_𝑋_*_2_* and 𝑏_𝑋_*_2_*_𝑌_*_2_*) and correlated genetic liabilities (𝑐𝑜𝑣𝐴2; **Figure 1D**). The proximal effects at wave 2 reflect causal influences that occur before wave 2 but are not captured by the distal (cross-lagged) effects, allowing us to detect causation even when the distal effects have been diminished by a long time interval.

Of note, we did not consider similar proximal effects at wave 1 for two reasons. First, testing the proximal effects at baseline is equivalent to fitting a separate cross-sectional model to wave 1 data, which adds unnecessary complexity to the CLPM. Second, the purely causal twin models (unidirectional and bidirectional) assume no confounding due to correlated genetic and/or environmental liabilities. Instead of applying cross-sectional causal models at wave 1, we freely estimated the correlated genetic and environmental liabilities at baseline, which were then transmitted to wave 2 via the autoregression of each construct, making it more plausible to assume no *additional* genetic or environmental confounding at wave 2.

#### Goodness-of-Fit and Model Selection

All structural equation modeling analyses were performed using *OpenMx* v2.21.3 (Neale et al., 2016) in R v4.4.0 (R Core Team, 2024). The models were fitted using weighted least squares estimation, and the goodness-of-fit was calculated as Browne’s pseudo chi-squared (𝜒*^2^*) statistic (Browne, 1984) and a corresponding *pseudo-*AIC [Akaike information criterion (Akaike, 1974)] (**Supplementary Methods**). Model comparison was performed using the Satorra-Bentler (SB) scaled-difference 𝜒*^2^*statistic (Satorra & Bentler, 2001), with degrees of freedom (DF) equal to the difference in the number of estimated parameters between the two models. In comparing nested models, we considered an SB 𝜒*^2^*p-value <0.05 to indicate a significantly worse fit of the more parsimonious model. When comparing models with an equal number of estimated parameters, SB 𝜒*^2^*is necessarily zero, in which case the model with the lowest pseudo-AIC (the fewest parameters) was considered the most parsimonious. Here, 𝛥𝐴𝐼𝐶 > *2* (for 𝐷𝐹 = *1*) indicated a significant worsening of model fit.

## Results

### MZ and DZ Twin Correlations

The within-trait and cross-trait twin correlations of *DepSx* and *CigDay* assessed at TEDS21 and TEDS26 are presented in **Table 2**. Averaged across the two repeated assessments, the MZ twin correlations were approximately twice the DZ correlations: 0.39 (SE = 0.02) vs. 0.20 (SE = 0.02) for *DepSx*, and 0.70 (SE = 0.04) vs. 0.37 (SE = 0.05) for *CigDay*, respectively. The AE model was consistently the most parsimonious across all univariate twin analyses (**Supplementary Tables S1, S2**), as previously reported for several behavioral outcomes in TEDS21 and COVID Study assessments (Rimfeld et al., 2022). Therefore, all CLPM models presented below were fitted subject to AE variance decomposition.

**Table 2.** Within-individual and Cross-twin Correlations at TEDS21 and TEDS26.

|  |  | Twin 1 |  |  |  | Twin 2 |  |  |  |
| --- | --- | --- | --- | --- | --- | --- | --- | --- | --- |
|  |  | *DepSx21* | *CigDay21* | *DepSx26* | *CigDay26* | *DepSx21* | *CigDay21* | *DepSx26* | *CigDay26* |
| Twin 1 | *DepSx21* | 1 | 0.24 (0.03) | 0.53 (0.02) | 0.21 (0.04) | 0.17 (0.02) | 0.09 (0.04) | 0.19 (0.02) | -0.03 (0.05) |
|  | *CigDay21* | 0.16 (0.05) | 1 | 0.18 (0.04) | 0.73 (0.03) | 0.12 (0.04) | 0.37 (0.06) | 0.03 (0.05) | 0.36 (0.07) |
|  | *DepSx26* | 0.56 (0.02) | 0.14 (0.06) | 1 | 0.23 (0.04) | 0.17 (0.02) | 0.12 (0.04) | 0.24 (0.02) | 0.03 (0.05) |
|  | *CigDay26* | 0.02 (0.06) | 0.71 (0.05) | 0.23 (0.05) | 1 | 0.01 (0.05) | 0.26 (0.07) | 0.03 (0.05) | 0.37 (0.08) |
| Twin 2 | *DepSx21* | 0.38 (0.02) | 0.17 (0.05) | 0.33 (0.03) | 0.09 (0.06) | 1 | 0.21 (0.03) | 0.51 (0.02) | 0.21 (0.04) |
|  | *CigDay21* | 0.06 (0.05) | 0.66 (0.05) | 0.07 (0.06) | 0.63 (0.06) | 0.23 (0.05) | 1 | 0.25 (0.04) | 0.77 (0.03) |
|  | *DepSx26* | 0.40 (0.03) | 0.23 (0.06) | 0.40 (0.03) | 0.20 (0.06) | 0.53 (0.02) | 0.20 (0.05) | 1 | 0.25 (0.04) |
|  | *CigDay26* | 0.11 (0.06) | 0.65 (0.06) | 0.21 (0.06) | 0.74 (0.05) | 0.23 (0.06) | 0.75 (0.04) | 0.20 (0.05) | 1 |

*Note.* The bottom off-diagonal triangle shows the correlations in MZ (monozygotic) twin pairs, while the top triangle shows the correlations in DZ (dizygotic) twin pairs. The values in parentheses indicate the standard error of the correlation.
Similar correlations across all six waves are shown in **Supplementary Table S3**.
**Color Legend.** Within-individual cross-trait cross-sectional correlation. Within-individual within-trait longitudinal correlation (i.e., autocorrelation). Within-individual cross-trait longitudinal (i.e., cross-lagged) correlation. Cross-twin within-trait cross-sectional correlation. Cross-twin cross-trait cross-sectional correlation. Cross-twin within-trait longitudinal correlation. Cross-twin cross-trait longitudinal (i.e., cross-lagged) correlation.

*DepSx* had a lower autocorrelation (0.53; SE = 0.02) than *CigDay* (0.74; SE = 0.04), indicating greater instability in *DepSx* than *CigDay*. For both variables, the cross-twin within-trait lagged correlations, e.g., between Twin 1’s *DepSx* at TEDS21 and Twin 2’s *DepSx* at TEDS26, were only marginally smaller than the corresponding cross-sectional twin correlations. The lagged MZ and DZ correlations were 0.37 (SE = 0.02) and 0.18 (SE = 0.02) for *DepSx*, and 0.64 (SE = 0.09) and 0.31 (SE = 0.10) for *CigDay*, respectively. As there was little difference between the cross-sectional and lagged twin correlations, the factors responsible for twin similarity at each assessment (i.e., the genetic factors) were likely stable across both constructs during the study period.

The average (within-individual) cross-sectional correlation between *DepSx* and *CigDay* was 0.22 (SE = 0.01). The average cross-twin cross-trait cross-sectional correlation was 0.16 (SE = 0.03) in MZ twins and 0.08 (SE = 0.02) in DZ twins, suggesting that correlated genetic liabilities contribute to the observed association between *DepSx* and *CigDay*. The within-individual cross-lagged correlations were similar in both directions: 0.19 (SE = 0.02) between *DepSx* at TEDS21 and *CigDay* at TEDS26, and 0.20 (SE = 0.02) between *CigDay* at TEDS21 and *DepSx* at TEDS26. However, the cross-twin cross-lagged correlations were higher from *CigDay* to *DepSx* (rMZ = 0.16 [SE = 0.04]; rDZ = 0.08 [SE = 0.03]) than from *DepSx* to *CigDay* (rMZ = 0.10 [SE = 0.04]; rDZ = – 0.01 [SE = 0.04]). **Supplementary Table S3** presents similar correlations of *DepSx* and *CigDay* across all six waves.

### Standard Twin CLPM: Lagged Effect of *CigDay* on *DepSx*

The standard two-wave twin CLPM (with phenotypic autoregressive processes; **Figure 1A**) indicated a unidirectional causal process, with *CigDay* predicting *DepSx* approximately four years later (standardized path coefficient, 𝛽 = 0.15; SE = 0.02), but not vice versa (𝛽 <0.001; SE = 0.03) (**Supplementary Table S4**; **Supplementary Figure S2**). That is, an increase of one standard deviation (SD) in the liability for *CigDay* predicted a 0.15 SD increase in the liability for depressive symptoms four years later, relative to one’s peers.

In the reduced, best-fitting model with a unidirectional *CigDay→DepSx* lagged association, the phenotypic autoregression 𝛽 was estimated to be 0.47 (SE = 0.01) for *DepSx* and 0.77 (SE = 0.02) for *CigDay* (**Table 3; Supplementary Table S5**; **Figure 2A**). Here, as the phenotypic autoregression mediates the temporal stability of genetic and environmental factors, the stability of these underlying influences is proportional to their contributions to the phenotypic variance. For instance, the genetic liability for *DepSx* accounted for approximately 36% of the stability of manifest *DepSx*, given that the latent genetic factor accounts for 36% of the variance in *DepSx*.

**Figure 2.**
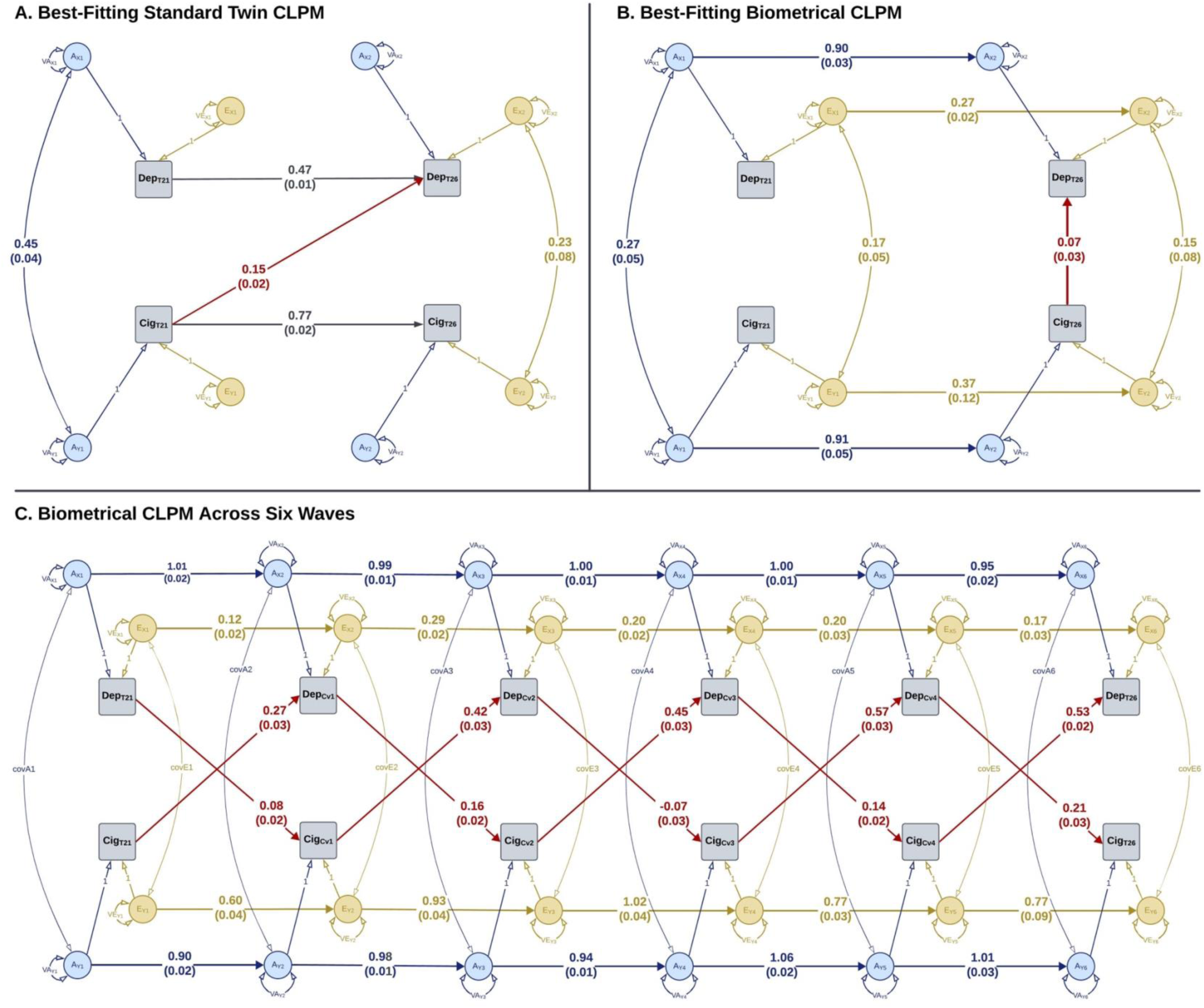
Causation Between DepSx and CigDay in Cross-Lagged Twin Models. **A.** Best-fitting standard twin CLPM fitted to TEDS21 and TEDS26 (time interval of approximately four years), with phenotypic autoregression and cross-lagged phenotypic paths (Model 10 in Table 3). **B. Best-titting Biometrical CLPM** fitted to TEDS21 and TEDS26 (time-interval of approximately four years), including latent genetic and environmental autoregression and phenotypic cross-sectional proximal effects (Model 18 in Table 3). This model with unidirectional proximal causation had only a marginally better fit than the bidirectional model (Model 14 in Table 3). **C. Biometrical CLPM** fitted to six waves (TEDS21, COVID Study Phase 1-4, and TEDS26) with variable intervals from two months to two years. Note. Standardized estimates of selected coefficients are shown, along with the standard error in parentheses. All parameter estimates are shown in **Supplementary Tables S5, S7, and S10**, respectively.

**Table 3.** Model Selection in (I.) the Standard Twin CLPM and (II.) the Biometrical CLPM.

| | Nr. | Model | EP | Browne's<br>$\chi^2$ | DF | AIC | Model<br>Comparison | ΔAIC | SB $\chi^2$ | ΔDF | SB $\chi^2$ p-value |
| --- | --- | --- | --- | --- | --- | --- | --- | --- | --- | --- | --- |
| **I. Standard Twin CLPM** |  |  |  |  |  |  |  |  |  |  |  |
| **A. Test Cross-Lagged Associations** |  |  |  |  |  |  |  |  |  |  |  |
|  | 1 | Cross-lagged associations | 20 | 444.27 | 68 | 484.27 |  |  |  |  |  |
|  | 2 | No cross-lagged associations | 18 | 489.27 | 70 | 525.27 | 2 v. 1 | 41.00 | 20.19 | 2 | <0.0001 |
|  | 3 | *Lagged CigDay → DepSx* | **19** | **444.30** | **69** | **482.30** | **3 v. 1** | **-1.98** | 0.01 | 1 | 0.9176 |
|  | 4 | *Lagged DepSx → CigDay* | 19 | 488.58 | 69 | 526.58 | 4 v. 1 | 42.31 | 19.45 | 1 | <0.0001 |
| **B. Test Shared Liabilities, in the Model with CigDay → DepSx effect (Model 3)** |  |  |  |  |  |  |  |  |  |  |  |
|  | 5 | No correlated A at TEDS21 | 18 | 513.30 | 70 | 549.30 | 5 v. 3 | 67.00 | 26.31 | 1 | <0.0001 |
|  | 6 | No correlated E at TEDS21 | 18 | 444.78 | 70 | 480.78 | 6 v. 3 | -1.51 | 0.18 | 1 | 0.6714 |
|  | 7 | No correlated A at TEDS26 | 18 | 444.97 | 70 | 480.97 | 7 v. 3 | -1.32 | 0.26 | 1 | 0.6107 |
|  | 8 | No correlated E at TEDS26 | 18 | 445.68 | 70 | 481.68 | 8 v. 3 | -0.62 | 0.53 | 1 | 0.4650 |
|  | 9 | No correlated AE at TEDS26 | 17 | 455.74 | 71 | 489.74 | 9 v. 3 | 7.44 | 4.79 | 2 | 0.0033 |
|  | 10 | **No correlated E at TEDS21; No correlated A at TEDS26** | **17** | **445.59** | **71** | **479.59** | **10 v. 3** | **-2.70** | 0.49 | 2 | 0.7833 |
| **II. Biometrical CLPM** |  |  |  |  |  |  |  |  |  |  |  |
| **A. Test Models of Within-Construct Stability in Twin CLPM with Cross-lagged Causation** |  |  |  |  |  |  |  |  |  |  |  |
|  | **11** | **AE autoregressions** | **22** | **297.09** | **66** | **341.09** |  |  |  |  |  |
|  | 1 | Phenotypic autoregressions | 20 | 444.27 | 68 | 484.27 | 1 v. 11 | 143.18 | 54.86 | 2 | <0.0001 |
| **B. Test Cross-Lagged Associations in CLPM with AE Autoregression (Model 11)** |  |  |  |  |  |  |  |  |  |  |  |
|  | 12 | Cross-lagged correlated AE Liabilities | 24 | 295.35 | 64 | 343.35 | 11 v. 12 | -2.25 | 0.66 | 2 | 0.7197 |
|  | 13 | **No cross-lagged associations** | **20** | **299.93** | **68** | **339.93** | **13 v. 12** | **-3.41** | 3.69 | 4 | 0.4500 |
|  |  |  |  |  |  |  | 13 v. 11 | -1.16 | 3.20 | 2 | 0.2023 |
| **C. Test Proximal Effects at TEDS26 in CLPM with AE Autoregression, but No Cross-Lagged Associations (Model 13)** |  |  |  |  |  |  |  |  |  |  |  |
|  | 13 | Cross-sectional correlated AE liabilities | 20 | 299.93 | 68 | 339.93 |  |  |  |  |  |
|  | 14 | Bidirectional Proximal Effects; No Correlated AE | 20 | 297.80 | 68 | 337.80 | 14 v. 13 | -2.13 | 0 | 0 | NA |
|  | 15 | Proximal *DepSx* → *CigDay*, and Correlated A | 20 | 301.93 | 68 | 341.93 | 15 v. 13 | 2.00 | 0 | 0 | NA |
|  | 16 | Proximal *DepSx* → *CigDay*, and Correlated E | 20 | 300.26 | 68 | 340.26 | 16 v. 13 | 0.33 | 0 | 0 | NA |
|  | 17 | Proximal *CigDay* → *DepSx*, and Correlated A | 20 | 298.26 | 68 | 338.26 | 17 v. 13 | -1.67 | 0 | 0 | NA |
|  | 18 | **Proximal *CigDay* → *DepSx*, and Correlated E** | **20** | **296.98** | **68** | **336.98** | **18 v. 13** | **-2.95** | 0 | 0 | NA |
|  |  |  |  |  |  |  | 18 v. 14 | -0.82 | 0 | 0 | NA |

*Note. EP = The number of estimated parameters. DF = Degrees of freedom. AIC = Akaike information criterion. SB $\chi^2$ = Satorra-Bentler scaled-difference $\chi^2$ statistic. SB $\chi^2 = 0$ in model comparisons with $\Delta DF = 0$ . The model shown in bold is the most parsimonious model within each section and is used as the starting point for the next section of tests.*

The covariance between *DepSx* and *CigDay* at baseline was attributable entirely to their correlated genetic liabilities (genetic correlation, 𝑟_𝐴_ = 0.45; SE = 0.04), which also contribute to the phenotypic covariance at TEDS26 via the phenotypic autoregression of each construct. The *additional* covariance between *DepSx* and *CigDay* at TEDS26 was due to the individual-specific environmental liabilities shared by the two phenotypes (environmental correlation, 𝑟_𝐸_ = 0.23; SE = 0.08).

### Biometrical CLPM: No Evidence of Lagged Effects Over Four Years

Compared to the Biometrical CLPM with distinct A and E autoregressions, the standard twin CLPM with phenotypic autoregression fit the data substantially worse (𝛥𝐴𝐼𝐶 = 143.18; SB 𝜒*^2^* p <0.0001; **Table 3**). In the Biometrical CLPM, the genetic factors had a much stronger autocorrelation than the environmental factors: *genetic* 𝛽_𝐴𝑅_ = 0.90 (SE = 0.03) and *environmental* 𝛽_𝐴𝑅_ = 0.27 (SE = 0.02) for *DepSx*, compared to the 0.47 autocorrelation for both in the standard CLPM (**Supplementary Table S6**; **Supplementary Figure S3**). Likewise, for *CigDay*, genetic factors had 𝛽_𝐴𝑅_ = 0.91 (SE = 0.05), and environmental factors had 𝛽_𝐴𝑅_ = 0.37 (SE = 0.12), in contrast to 0.77 for both in the previous model. Based on this model, the stability of the genetic liability was estimated to account for 65.6% (SE = 2.8%) of the phenotypic stability in *DepSx* and 86.2% (SE = 4.9%) in *CigDay*, with the remaining attributable to the environmental factors.

Further examining the sources of cross-lagged associations, the Biometrical CLPM with prospective *phenotypic* associations was more parsimonious than the one with correlated A and E liabilities (𝛥𝐴𝐼𝐶 = –2.25; SB 𝜒*^2^* p = 0.7197). However, compared to the standard CLPM above, the prospective association of *CigDay* with *DepSx* was attenuated considerably (𝛽 = 0.04; SE = 0.03). The lagged association of *DepSx* with *CigDay* was close to zero (𝛽 = 0.01; SE = 0.03), as before. Expectedly, the “null-hypothesis” model with no cross-lagged associations fit the data best: 𝛥𝐴𝐼𝐶 = –1.16; SB 𝜒*^2^* p = 0.2023. Therefore, contrary to the inference based on the standard CLPM, we rejected the hypothesis of lagged causal effects of *CigDay* on *DepSx*.

### Proximal Direction-of-Causation: Potential Short-Term Effects

As there was no evidence of distal effects in the Biometrical CLPM over a four-year interval, we examined proximal causal effects at TEDS26 using different specifications of *Biometrical CLPM-DoC* (**Table 3**). Of these models, the one with a unidirectional *CigDay → DepSx* proximal effect (𝛽 = 0.07; SE = 0.03) and correlated environmental liabilities (𝑟_𝐸_ = 0.15; SE = 0.08) fit the data best (**Supplementary Table S7**; **Figure 2B**). However, this model had only a marginally better fit than the one with bidirectional proximal effects, but no genetic or environmental correlated liabilities (𝛥𝐴𝐼𝐶 = −*0*.*82*), with 𝛽_𝐷𝑒𝑝𝑆𝑥 → 𝐶𝑖𝑔𝐷𝑎𝑦_ = 0.04 (SE = 0.03) and 𝛽_𝐶𝑖𝑔𝐷𝑎𝑦 → 𝐷𝑒𝑝𝑆𝑥_ = 0.08 (SE = 0.03) (**Supplementary Table S8**; **Supplementary Figure S4**). Taken together, these models indicate possible bidirectional *short-term* causal influences between *CigDay* and *DepSx*, with a relatively stronger effect of *CigDay* on *DepSx* than vice versa.

Notably, in contrast to the standard CLPM, the covariance between *CigDay* and *DepSx* at baseline was attributable to both correlated genetic (63.8%) and environmental (36.2%) factors in the Biometrical CLPM, with 𝑟_𝐴_ = 0.27 (SE = 0.05) an 𝑟_𝐸_ = 0.17 (SE = 0.05).

### Bidirectional Lagged Effects Over Shorter Intervals

We fitted the standard and Biometrical CLPMs to all six waves to examine potential bidirectional lagged/distal effects over shorter intervals. As before, the standard twin phenotypic CLPM fit the data considerably worse than the Biometrical CLPM with its independent and distinct genetic and environmental autoregressive processes (𝛥𝐴𝐼𝐶 = 947.94; SB 𝜒*^2^* p <0.0001; **Table S9**). In the Biometrical CLPM, nested models with either no cross-lagged paths or unidirectional lagged paths in either direction had significantly worse fit than the full model. The cross-lagged prospective associations varied across study waves, which may be due to developmental changes, contextual moderators (e.g., the COVID-19 crisis), and varying time intervals ranging from approximately two months between COVID Study Phases 1 and 2 to two years between TEDS21 and COVID Phase 1. The average cross-lagged estimates were: 𝛽_𝐶𝑖𝑔𝐷𝑎𝑦 → 𝐷𝑒𝑝𝑆𝑥_ = 0.45 (SE = 0.2), and 𝛽_𝐷𝑒𝑝𝑆𝑥 → 𝐶𝑖𝑔𝐷𝑎𝑦_ = 0.11 (SE = 0.05; **Supplementary Table S10**; **Figure 2C**). These findings support bidirectional causal effects between *CigDay* and *DepSx* persisting for at least two years, with a much stronger effect of *CigDay* on *DepSx* than vice versa, consistent with the proximal effect estimates four years apart.

## Discussion

Our novel biometrical cross-lagged panel model revealed bidirectional causal effects between smoking quantity and depressive symptoms in young adults. While the effects persisted for at least two years, they were not detectable after four years. The bidirectional causation, with positive coefficients in both directions, suggests a mutually reinforcing feedback loop that may contribute to the maintenance of smoking behaviors and dependence (Leventhal & Zvolensky, 2015; Mathew et al., 2017). The potential short-term effects of depressive symptoms on smoking quantity support the self-medication theory (Audrain-McGovern et al., 2009), which entails that the immediate (but transient) mood-elevating effects of smoking (Hedeker et al., 2009) motivate individuals to smoke more cigarettes to alleviate their depressive symptoms. Long-term cigarette smoking may, in turn, increase the vulnerability to depression through the chronic effects of nicotine exposure on neurobiological (Danielson et al., 2011; Rao, 2006), cardiovascular (Ezzati & Lopez, 2003), and inflammatory (Shiels et al., 2014) processes involved in depression etiology (Beydoun et al., 2020; Hare et al., 2014). This feedback loop is consistent with the potential role of negative affect (depressed mood) in nicotine dependence (Parrott, 1998). While smoking decreases negative affect temporarily, higher nicotine dependence (as indicated by heavier smoking) results in a rebound increase in negative affect when the blood nicotine levels drop between instances of smoking. Relief of these aversive symptoms upon smoking motivates continued use to maintain nicotine levels above the withdrawal threshold (Parrott, 1998). Such symptom relief has also been observed when modeling a feedback loop between a maintenance factor of the Fagerström Test for Nicotine Dependence and its Cigarettes-per-day item (Verhulst et al., 2021).

In addition to reciprocal causation, there was evidence of genetic and environmental factors that increase the susceptibility to both smoking quantity and depressive symptoms. The correlated genetic liability may reflect neuropsychological traits, such as behavioral disinhibition, potentially contributing to both internalizing (e.g., depression) and externalizing (e.g., substance use) psychopathology (Nigg, 2017). Likewise, gene-prioritization methods applied to genome-wide association studies of depression and smoking have revealed shared biological pathways involving neurotransmitter systems, e.g., membrane potential, GABA-receptor activity, and endocannabinoid signaling pathways (Yao et al., 2021). Notably, the non-causal association between smoking quantity and depressive symptoms in mid- to late-twenties (TEDS26), beyond the shared risk factors in early twenties (TEDS21), was primarily due to environmental factors. These additional correlated environmental factors may reflect the stressors and protective factors associated with many possible life changes in the later twenties, including the transition from student to working life, moving out of the parents’ home, entering a long-term romantic partnership or marriage, and becoming a parent (Arnett, 2024). As this cohort was impacted by the COVID-19 pandemic in their mid-twenties, the pandemic and the ensuing lockdowns may have added to the environmental factors shared between depressive symptoms and smoking quantity (Douglas et al., 2020). Despite the increased environmental confounding, the proportions of phenotypic variance in depressive symptoms and smoking quantity attributable to environmental factors remained stable over time (**Supplementary Table S2**), consistent with several other mental health outcomes in this cohort (Rimfeld et al., 2022).

The Biometrical CLPM provided three novel insights over the standard twin CLPM. First, the autocorrelations of the genetic liabilities for smoking quantity and depressive symptoms are substantially larger than those of the environmental influences. Notably, accounting for these autocorrelation differences attenuated the cross-lagged causal estimates in the Biometrical CLPM compared to the standard CLPM. Second, the Biometrical CLPM included potential cross-lagged confounding due to shared genetic and environmental influences, which were assumed absent in the standard CLPM. However, we did not find evidence of cross-lagged confounding, nor causal influences, between smoking and depressive symptoms assessed roughly four years apart. Third, the Biometrical CLPM incorporated cross-sectional proximal effects, which suggested short-term bidirectional causal effects, though the effect of smoking quantity on depressive symptoms was stronger. Finally, the Biometrical CLPM fitted to six study waves with shorter intervals (from two months to two years) revealed significant bidirectional lagged effects, consistent with the proximal effects estimated at a four-year interval.

To our knowledge, the Biometrical CLPM is the first *multi-phenotype* developmental twin model, advancing the long-standing tradition of *single-phenotype* models (e.g., (Boomsma & Molenaar, 1987; Dolan et al., 2014; Eaves et al., 1986; McArdle, 1986) to help address causality and correlated liabilities between co-developing constructs in longitudinal twin data. This model includes autoregressive processes of stability, but not a random intercept, which represents the trait-like time-invariant stability of a construct. The random-intercept CLPM (RI-CLPM) is better suited for identifying the *within-individual* cross-lagged effects of short-term *state-like deviations* from an individual’s long-term average trait (Hamaker et al., 2015). Meanwhile, the cross-lagged effects in CLPM are interpretable as the effects of the *rank order* (relative to the population) of one variable on another (Orth et al., 2021). Thus, the causal effects identified in this study may help explain partly why individuals who smoke more heavily than their peers experience, on average, higher depressive symptoms than their peers. As such, CLPM remains valuable for understanding the *between-individual* differences at the population level (Lüdtke & Robitzsch, 2021). Future extensions of the Biometrical CLPM to RI-CLPM may build on another developmental twin model that includes separate genetic and environmental random intercepts and autoregressive processes (Eaves et al., 1986).

In this study, the cross-sectional proximal causal effects were estimated based on the cross-twin cross-trait correlations (Heath et al., 1993), which required assumptions of no background confounding due to one or more cross-sectional covariance parameters. Alternatively, previous studies have estimated cross-sectional causal paths and the possible covariance parameters by integrating IVs in cross-sectional twin models (Castro-de-Araujo et al., 2022; Minică et al., 2018; Singh et al., 2025) and CLPM (Singh et al., 2024). Building on these approaches, our ongoing work further integrates IVs into the presented Biometrical CLPM, as outlined in our pre-registration. However, these models require thorough theoretical exposition and careful empirical testing of the IV assumptions (Labrecque & Swanson, 2018), which are beyond the present scope.

When interpreting these results, it should be noted that the assessment of current smoking quantity was limited to those who had ever smoked a cigarette, since smoking initiation and quantity (or degree of dependence) have distinct relationships with depression (Edwards & Kendler, 2012; Kendler et al., 1999). We did not include smoking initiation as an additional phenotype in the models because, once an individual has initiated smoking, lifetime initiation is temporally invariant, making it unsuitable for longitudinal analyses using CLPM. Moreover, the sources of covariance between smoking quantity and depressive symptoms may differ by sex, which we did not test in this study. That said, previous sex-stratified analyses of substance dependence and depression in young adults found no significant sex differences in the cross-lagged associations (Marmorstein et al., 2010). Also, we found no evidence of sex differences in smoking quantity, consistent with previous analyses of two US-based datasets (Bares et al., 2015; Do et al., 2015). Lastly, we examined data from a community-based cohort, wherein most participants endorsed few or no depressive symptoms, and only a small fraction reported heavy smoking. The generalizability of our findings to individuals with a clinical diagnosis of MDD and/or nicotine dependence requires further research. Likewise, these analyses were limited to conventional *cigarette* smoking. As e-cigarette use is becoming an increasingly prevalent form of tobacco use (Jackson et al., 2024), the generalizability of the current findings to e-cigarette use has yet to be examined.

These limitations notwithstanding, this study sheds light on bidirectional causation between smoking quantity and depressive symptoms in young adults, as well as their shared genetic and environmental liabilities. Results across models suggest that prospective causal influences remain appreciable for at least two years but dissipate over longer intervals. Through these analyses, this study presents a series of novel cross-lagged twin models including separate genetic and environmental developmental processes, comparisons of cross-lagged confounding and causation, and tests of proximal short-term causal effects. These models may enable more nuanced and robust causal inference in longitudinal twin studies, which, in turn, may inform public health measures.

## Data Availability

TEDS data are available upon request. Procedures are described here: https://www.teds.ac.uk/researchers/teds-data-access-policy.

## Code Availability

Code for fitting the Biometrical CLPM models is available on GitHub, here: https://github.com/singh-madhur/BiometricalCLPM.

## Author Notes

### CRediT Statement

**Madhurbain Singh**: Conceptualization, Formal analysis, Methodology, Software, Visualization, Writing—original draft, Writing—review and editing. **Michael D. Hunter**: Software, Writing—review and editing. **Elham Assary**: Data curation, Writing—review and editing. **Brad Verhulst**: Methodology, Writing—review and editing. **Roseann E. Peterson:** Funding acquisition, Writing—review and editing. **Hermine H. M. Maes**: Methodology, Writing—review and editing. **Conor V. Dolan**: Conceptualization, Methodology, Project administration, Software, Supervision, Writing—review and editing. **Thalia C. Eley**: Conceptualization, Data curation, Project administration, Resources, Supervision, Writing—review and editing. **Michael C. Neale**: Conceptualization, Funding acquisition, Methodology, Project administration, Resources, Software, Supervision, Writing—review and editing.

## Acknowledgements

This work was supported by the U.S. National Institutes of Health grants R01DA049867 (MCN, MS, CVD, HHMM) and R01MH125938 (REP, MS).

We gratefully acknowledge the ongoing contribution of the participants in the Twins Early Development Study (TEDS) and their families. TEDS is supported by the UK Medical Research Council (MR/V012878/1 and previously MR/M021475/1).

## Notes

### Competing Interest Statement

The authors have declared no competing interest.

### Author Declarations

TEDS is approved by the King's College London Research Ethics Committee (References: PNM/09/10-104 and HR/DP-20/21-22060).

