## Supplementary Information for "Causation Between Smoking Quantity and Depressive Symptoms in Young Adults: Evidence From Novel Cross-Lagged Twin Models"

**SUPPLEMENTARY MATERIAL**

Madhurbain Singh, Michael D. Hunter, Elham Assary, Brad Verhulst, Roseann E. Peterson, Hermine H. M. Maes, Conor V. Dolan*, Thalia C. Eley*, and Michael C. Neale*

*Joint last authors

### Supplementary Note: Deviations from the Preregistration

The work presented in this manuscript is part of our pre-registered project examining the shared etiology and causation between depression, anxiety, and substance use in young adults: https://doi.org/10.17605/OSF.IO/K47BF; pre-registered on June 11, 2024. The overall project aims to examine the causal effects and shared genetic and environmental liabilities between internalizing psychopathology (depression or anxiety symptoms) and substance use (cigarette smoking or alcohol use), by fitting pairwise models between one of the internalizing symptom scales and one of the substance use outcome measures. The current manuscript focuses on methodological issues and proposed solutions, using one such pair of constructs as an example: depressive symptoms and smoking quantity (cigarettes per day).

In line with the preregistered hypotheses, the current manuscript sought to test the following hypotheses:

1. The cross-sectional covariance between depressive symptoms and cigarettes per day in young adults can be partitioned into:
    a. Small, positive covariance between latent additive genetic factors,
    b. Small, positive covariance between latent individual-specific environmental factors,
    c. Small, positive causal effects of depressive symptoms on cigarettes per day, and
    d. Small, positive causal effects of cigarettes per day on depressive symptoms.
2. The longitudinal covariance between repeated measures of depressive symptoms and cigarettes per day can be partitioned into:
    a. Small, positive, cross-lagged bidirectional causal effects,
    b. Small, positive, cross-lagged cross-trait influences between latent additive genetic factors, and
    c. Small, positive, cross-lagged cross-trait influences between latent individual-specific environmental factors.

In our preregistration, we also hypothesized the role of covariance between latent familial environmental factors underlying the two constructs. However, our preliminary univariate twin models did not support the role of familial environmental factors in the variance of either construct. Therefore, latent familial environmental factors were not included in the twin cross-lagged panel models (CLPMs).

In our preregistered analysis plan, we proposed to test cross-sectional ("proximal") causal effects in the cross-lagged twin models by introducing instrumental variables (IVs) into the model (Bollen, 2012; Maydeu-Olivares et al., 2020), essentially developing a twin version of the previously developed IV-CLPM (Singh et al., 2024) for data from unrelated individuals. However, before using IVs in the model, it is vital to introduce the assumptions of a valid IV and perform a thorough empirical examination of the validity of these assumptions in the observed data (Labrecque & Swanson, 2018). The current manuscript addresses three potential limitations of the standard twin CLPM and proposes possible methodological solutions. Therefore, further introduction of IVs and a twin IV-CLPM was deemed outside the current scope. The hypothesized cross-sectional causal paths were instead examined by integrating the twin direction-of-causation models (Heath et al., 1993; Neale & Cardon, 1992) into the CLPM. The proposed integration of IVs in twin CLPM will be presented in a separate manuscript.

Lastly, we proposed to use full information maximum likelihood (FIML) to fit the models, which requires using raw data. However, due to data access limitations, the analyses were performed in a two-step approach, where we first obtained the summary statistics and their associated weights from the raw data. As summary statistics do not contain individual-level information, these data objects could be shared across collaborators and institutes. The models were then fitted to the summary statistics using a full weighted least squares (WLS) estimator.

### Supplementary Methods

#### Study Sample

Over 13,000 twin pairs were assessed at the baseline TEDS assessment when the twins were 18 months old. During young adulthood, over 8,000 twin pairs were contactable and invited to participate across six repeated assessments over five years (2018–22) (Lockhart et al., 2023; Rimfeld et al., 2022; Rimfeld et al., 2019). Two of these assessments were core TEDS study waves, conducted at an interval of 3.5 to 4.5 years: "TEDS-21" assessment in 2018, with responses from 8,005 individuals aged 21–26 (median 22.9 years) (Rimfeld et al., 2019), and "TEDS-26" assessment in 2021–22, with responses from 7,927 individuals at ages 25–29 (median 26.4 years). The four *COVID Study* assessments were completed in April, July, October 2020, and March 2021, approximately 1, 4, 7, and 11 months after the first national lockdown in Britain in March 2020. The total number of participants in the COVID Study was 4596 in Phase 1, 3800 in Phase 2, 3470 in Phase 3, and 3744 in Phase 4.

In total, we examined data from 10,034 participants with at least one assessment of smoking quantity (N = 6,326) or depressive symptoms (N = 10,003) across the six waves. The study sample included more females (6,177; 61.6%) than males (3,857; 38.4%). The participants were clustered into 5,922 twin pairs, of which 4,112 were complete twin pairs, including 1,042 MZ female, 531 MZ male, 903 DZ female, 426 DZ male, and 1,210 DZ opposite-sex twin pairs. The two-wave models fitted to TEDS-21 and TEDS-26 data included data from 9,606 individuals (62.0% females), of which 3,875 were complete twin pairs (994 MZ female, 507 MZ male, 857 DZ female, 397 DZ male, and 1,120 DZ opposite-sex twin pairs).

#### Smoking Quantity: Cigarettes per Day (*CigDay*)

At each study wave, the participants were first asked if they had ever smoked a cigarette. Those who reported ever smoking were asked if they were currently smoking and, if so, how many cigarettes a day they smoked (*CigDay*), with response options "10 or less," "11–20," and "21–30," and "31 or more." The responses were missing for individuals who had never smoked a cigarette, as well as for those who had initiated smoking but were not smoking at the time of assessment. Since smoking initiation and quantity have distinct genetic and environmental liabilities (Bares et al., 2015; Do et al., 2015), we re-coded the missing *CigDay* values as zero for only those who had initiated smoking but were not currently smoking. Very few individuals reported smoking more than 20 cigarettes per day ("21–30" or "31 or more"), e.g., 11 individuals in COVID Phase-4 to 36 in TEDS-21. Therefore, we merged the top three levels of the scale to obtain three levels of current *CigDay*: *None*, *Light* (up to 10 cigarettes per day), and *Heavy* (more than 10 cigarettes per day). This ordinal variable was examined using the liability threshold model, assuming a normally distributed latent liability (Verhulst & Neale, 2021), with mean and variance estimated freely at each wave, while the thresholds were fixed at arbitrary values of 0.5 and 1.5 (Mehta et al., 2004).

There were minimal sex differences in *CigDay*, with 6% of females and 7% of males reporting *Heavy* current smoking, and 19% and 17%, respectively, reporting *Light* current smoking across study waves. We applied ordered probit regression to all *CigDay* observations using the `polr()` function from the `MASS` R package (Venables & Ripley, 2002), which showed small, non-significant associations with age ($b = -0.007, SE = 0.007$) and sex ($b = -0.018, SE = 0.022$ [males compared to females]). Note that this regression model assumed independent observations, making the standard errors smaller than they should be. In a model accounting for the clustering of observations within individuals and twin pairs, the standard errors would be larger, further indicating no evidence of associations of *CigDay* with age and sex.

#### Depressive Symptoms (*DepSx*)

In TEDS-21 and COVID Study waves, recent depressive symptoms were assessed using the 8-item *Short Mood and Feelings Questionnaire (SMFQ)* (Angold et al., 1995). MFQ assesses the respondent's depressive symptoms over the preceding two weeks, scoring each symptom on a three-point ordinal scale of "Not True," "Quite True," and "Very True." The eight items included in SMFQ inquired if the respondent "felt miserable or unhappy," "felt so tired [they] just sat around and did nothing," "felt very restless," "cried a lot," "found it hard to think properly or concentrate," "hated [themself]," "felt lonely," "thought [they] could never be as good as other people." The response to each item was coded as 0, 1, and 2, respectively, and combined into a sum score (range 0–16) if a participant answered at least four (50%) of the items. In TEDS-26, the full 13-item MFQ was assessed. However, to obtain consistent repeated measures, we restricted the TEDS-26 responses to the eight SMFQ items assessed at earlier waves, obtaining sum scores ranging from 0 to 16.

The SMFQ sum scores (**Table 1**) were right-skewed, as is expected in psychopathology scales. To approximate a normal distribution, we first regressed the SMFQ sum scores (a total of 30,599 observations across all participants and study waves) on age ($b = 0.076$; $SE = 0.014$) and sex ($b = -1.213$; $SE = 0.048$). A rank-based inverse-normal transformation was then applied to the residuals from this regression model (skew 0.95, kurtosis 0.21), using the `RankNorm()` function from the `RNOmni` R package (McCaw, 2023). Like *CigDay*, this transformation was based on the assumption that the observed depressive symptom scores, conditional on age and sex, are a function of an underlying liability normally distributed in the population. Here, the residuals were first ranked from smallest to largest, and then the ranks were converted into quantiles (cumulative probabilities). Finally, the quantiles were transformed into Z-scores using the inverse of the standard normal cumulative distribution function (i.e., the probit function).

#### Standard Twin CLPM

We fitted the standard two-wave twin CLPM {Burt, S. Alexandra et al., 2005} to TEDS-21 (wave 1) and TEDS-26 (wave 2) data (**Figure 1A**), including cross-lagged causal paths between *DepSx* and *CigDay* ($b_{Y2X1}$ and $b_{X2Y1}$) and phenotypic autoregression within each construct ($arX$ and $arY$). To control for the clustering of data within twin pairs in this model, we applied biometrical (ACE) variance decomposition to the phenotypic variances and covariance on each occasion, estimating the genetic and environmental sources of the variance of each phenotype (e.g., $VA_{X1}$, $VC_{X1}$, and $VE_{X1}$ for *DepSx* at wave 1) and the cross-sectional covariance between phenotypes (e.g., $covA1$, $covC1$, and $covE1$ between *DepSx* and *CigDay* at wave 1). As in the classical twin design (Neale & Cardon, 1992), the cross-twin correlation between respective additive genetic (A) factors was fixed at 1 in MZ and 0.5 in DZ twin pairs. The cross-twin correlation between the familial environmental (C) factors was fixed at 1 in both twin pairs. The phenotypic cross-lagged path coefficients in this model are interpreted as potential causal effects, consistent with Granger causality (Granger, 1969). To test the bidirectional causal hypothesis, a "null" model was fitted with the phenotypic cross-lagged path coefficients fixed at zero. If the null model fits the data significantly worse than the full CLPM, we can reject the null hypothesis and infer potential causation. Finally, the genetic and environmental sources of cross-sectional covariance were tested by fixing these path coefficients to zero and comparing the goodness-of-fit statistics across models. The measures of goodness-of-fit and model comparison used in this study are described further below.

#### Goodness of Fit and Model Selection

All structural equation modeling (SEM) analyses were performed using the `OpenMx` package, v2.21.3 (Neale et al., 2016), in R v4.4.0 (R Core Team, 2024). All models were fitted using the full weighted least squares (WLS) estimator in a two-step process. In Step 1, we obtained summary statistics and the associated weights by applying the `omxAugmentDataWithWLSSummary()` function to the raw data. In Step 2, the model was fitted to the Step-1 summary object using the `mxFitFunctionWLS()` estimator. Both steps used the argument `type="WLS"`. In WLS models, the overall model-fit statistic calculated by `OpenMx` is the pseudo chi-squared ($\chi^2$) statistic by Browne (Browne, 1984). This pseudo-$\chi^2$ is also converted into a *pseudo*-AIC [Akaike information criterion (Akaike, 1974)] as: *pseudo*-$\chi^2 + (2 \times k)$, where $k$ is the number of estimated parameters. For comparing models fitted with WLS, `OpenMx`'s `mxCompare()` function provides the Satorra-Bentler (SB) scaled-difference $\chi^2$ statistic (Satorra & Bentler, 2001), with its degrees of freedom equaling the difference in the number of estimated parameters in the two models.

### Supplementary Tables

#### Table S1

*Model Fit Statistics in Univariate Twin Models*

| | | Base Model | Comparison Model | EP | -2LL | DF | AIC | Δ-2LL | ΔDF | p-value |
|---|---|---|---|---|---|---|---|---|---|---|
| ***CigDay*** | TEDS-21 | ACE | | 7 | 5,918.31 | 4415 | 5,932.31 | | | |
| | | ACE | AE | 6 | 5,918.49 | 4416 | 5,930.49 | 0.19 | 1 | 0.6660 |
| | COVID-1 | ACE | | 7 | 3,142.77 | 2143 | 3,156.77 | | | |
| | | ACE | AE | 6 | 3,143.16 | 2144 | 3,155.16 | 0.39 | 1 | 0.5332 |
| | COVID-2 | ACE | | 7 | 2,629.55 | 1735 | 2,643.55 | | | |
| | | ACE | AE | 6 | 2,634.08 | 1736 | 2,646.08 | 4.53 | 1 | 0.0333 |
| | COVID-3 | ACE | | 7 | 2,267.59 | 1538 | 2,281.59 | | | |
| | | ACE | AE | 6 | 2,267.61 | 1539 | 2,279.61 | 0.02 | 1 | 0.8811 |
| | COVID-4 | ACE | | 7 | 2,276.80 | 1654 | 2,290.80 | | | |
| | | ACE | AE | 6 | 2,281.51 | 1655 | 2,293.51 | 4.71 | 1 | 0.0299 |
| | TEDS-26 | ACE | | 7 | 4,596.58 | 4012 | 4,610.58 | | | |
| | | ACE | AE | 6 | 4,596.58 | 4013 | 4,608.58 | 0.00 | 1 | 0.9650 |
| ***DepSx*** | TEDS-21 | ACE | | 4 | 20,165.29 | 7700 | 20,173.29 | | | |
| | | ACE | AE | 3 | 20,165.67 | 7701 | 20,171.67 | 0.38 | 1 | 0.5390 |
| | COVID-1 | ACE | | 4 | 12,056.96 | 4468 | 12,064.96 | | | |
| | | ACE | AE | 3 | 12,058.33 | 4469 | 12,064.33 | 1.37 | 1 | 0.2414 |
| | COVID-2 | ACE | | 4 | 10,292.68 | 3711 | 10,300.68 | | | |
| | | ACE | AE | 3 | 10,293.26 | 3712 | 10,299.26 | 0.58 | 1 | 0.4450 |
| | COVID-3 | ACE | | 4 | 9,666.07 | 3371 | 9,674.07 | | | |

| | | | | | | | | | | |
|---|---|---|---|---|---|---|---|---|---|---|
| | | ACE | AE | 3 | 9,666.12 | 3372 | 9,672.12 | 0.06 | 1 | 0.8117 |
| | COVID-4 | ACE | | 4 | 10,343.48 | 3574 | 10,351.48 | | | |
| | | ACE | AE | 3 | 10,343.91 | 3575 | 10,349.91 | 0.43 | 1 | 0.5123 |
| | TEDS-26 | ACE | | 4 | 23,027.62 | 7751 | 23,035.62 | | | |
| | | ACE | AE | 3 | 23,031.49 | 7752 | 23,037.49 | 3.87 | 1 | 0.0492 |

**Table S2**

*Estimates of A and E Variance Components in Univariate Twin Models*

| | | Parameter | 95% L.B. | Estimate | 95% U.B. |
|---|---|---|---|---|---|
| ***CigDay*** | TEDS-21 | **A** | 0.58 | **0.68** | 0.75 |
| | | **E** | 0.31 | **0.32** | 0.42 |
| | COVID-1 | **A** | 0.49 | **0.65** | 0.76 |
| | | **E** | 0.24 | **0.35** | 0.51 |
| | COVID-2 | **A** | 0.29 | **0.48** | 0.64 |
| | | **E** | 0.36 | **0.52** | 0.71 |
| | COVID-3 | **A** | 0.52 | **0.69** | 0.82 |
| | | **E** | 0.18 | **0.31** | 0.48 |
| | COVID-4 | **A** | 0.50 | **0.67** | 0.79 |
| | | **E** | 0.21 | **0.33** | 0.50 |
| | TEDS-26 | **A** | 0.64 | **0.74** | 0.82 |
| | | **E** | 0.18 | **0.26** | 0.36 |
| ***DepSx*** | TEDS-21 | **A** | 0.32 | **0.36** | 0.40 |
| | | **E** | 0.60 | **0.64** | 0.68 |
| | COVID-1 | **A** | 0.34 | **0.40** | 0.45 |
| | | **E** | 0.55 | **0.60** | 0.66 |
| | COVID-2 | **A** | 0.31 | **0.38** | 0.44 |
| | | **E** | 0.56 | **0.62** | 0.69 |
| | COVID-3 | **A** | 0.30 | **0.37** | 0.43 |
| | | **E** | 0.57 | **0.63** | 0.70 |
| | COVID-4 | **A** | 0.32 | **0.39** | 0.45 |
| | | **E** | 0.55 | **0.61** | 0.68 |

| | | Parameter | 95% L.B. | Estimate | 95% U.B. |
|---|---|---|---|---|---|
| | TEDS-26 | **A** | 0.36 | **0.40** | 0.44 |
| | | **E** | 0.56 | **0.60** | 0.64 |
| | | **Unstandardized Estimates** | | | |
| ***DepSx*** **(Unstandardized)** | TEDS-21 | **A** | 0.26 | **0.30** | 0.34 |
| | | **E** | 0.49 | **0.53** | 0.56 |
| | COVID-1 | **A** | 0.30 | **0.35** | 0.41 |
| | | **E** | 0.49 | **0.54** | 0.59 |
| | COVID-2 | **A** | 0.29 | **0.36** | 0.43 |
| | | **E** | 0.54 | **0.60** | 0.66 |
| | COVID-3 | **A** | 0.31 | **0.39** | 0.47 |
| | | **E** | 0.60 | **0.66** | 0.74 |
| | COVID-4 | **A** | 0.34 | **0.42** | 0.50 |
| | | **E** | 0.60 | **0.66** | 0.74 |
| | TEDS-26 | **A** | 0.42 | **0.48** | 0.53 |
| | | **E** | 0.66 | **0.70** | 0.75 |

**Table S3**

*Within-individual and Cross-twin Correlations at TEDS-21, COVID Study Phase 1–4, and TEDS-26*

| | | Twin 1 | | | | | | | | | | | | Twin 2 | | | | | | | | | | | |
|---|---|---|---|---|---|---|---|---|---|---|---|---|---|---|---|---|---|---|---|---|---|---|---|---|---|
| | | DepT1 | CigT1 | DepT2 | CigT2 | DepT3 | CigT3 | DepT4 | CigT4 | DepT5 | CigT5 | DepT6 | CigT6 | DepT1 | CigT1 | DepT2 | CigT2 | DepT3 | CigT3 | DepT4 | CigT4 | DepT5 | CigT5 | DepT6 | CigT6 |
| Twin 1 | Dep T1 | 1 | 0.24 (0.03) | 0.56 (0.02) | 0.22 (0.05) | 0.55 (0.02) | 0.24 (0.05) | 0.53 (0.02) | 0.16 (0.06) | 0.54 (0.02) | 0.23 (0.05) | 0.53 (0.02) | 0.2 (0.04) | 0.17 (0.02) | 0.09 (0.04) | 0.18 (0.03) | 0.1 (0.06) | 0.16 (0.03) | 0.14 (0.06) | 0.16 (0.04) | 0.2 (0.06) | 0.21 (0.03) | 0.1 (0.07) | 0.19 (0.02) | -0.02 (0.05) |
| | Cig T1 | 0.16 (0.05) | 1 | 0.15 (0.05) | 0.8 (0.03) | 0.24 (0.05) | 0.8 (0.03) | 0.21 (0.06) | 0.71 (0.05) | 0.28 (0.05) | 0.79 (0.03) | 0.18 (0.04) | 0.74 (0.03) | 0.12 (0.04) | 0.37 (0.06) | 0.13 (0.06) | 0.4 (0.08) | 0.12 (0.06) | 0.35 (0.09) | 0.19 (0.07) | 0.33 (0.1) | 0.14 (0.07) | 0.48 (0.09) | 0.03 (0.05) | 0.36 (0.07) |
| | Dep T2 | 0.55 (0.03) | 0.09 (0.07) | 1 | 0.15 (0.05) | 0.72 (0.02) | 0.19 (0.06) | 0.68 (0.02) | 0.11 (0.06) | 0.62 (0.02) | 0.17 (0.06) | 0.58 (0.02) | 0.13 (0.06) | 0.16 (0.03) | 0.06 (0.05) | 0.16 (0.04) | 0.04 (0.07) | 0.22 (0.04) | 0.04 (0.07) | 0.2 (0.04) | 0.14 (0.07) | 0.2 (0.04) | 0.07 (0.08) | 0.16 (0.03) | -0.04 (0.07) |
| | Cig T2 | 0.13 (0.07) | 0.78 (0.04) | 0.1 (0.07) | 1 | 0.18 (0.06) | 0.93 (0.01) | 0.2 (0.07) | 0.88 (0.02) | 0.25 (0.06) | 0.89 (0.02) | 0.18 (0.05) | 0.81 (0.03) | 0.1 (0.06) | 0.3 (0.08) | 0.14 (0.07) | 0.37 (0.1) | 0.11 (0.08) | 0.35 (0.1) | 0.16 (0.08) | 0.35 (0.11) | 0.09 (0.08) | 0.36 (0.12) | 0.06 (0.06) | 0.24 (0.1) |
| | Dep T3 | 0.57 (0.03) | 0.08 (0.08) | 0.7 (0.02) | 0.07 (0.08) | 1 | 0.23 (0.05) | 0.71 (0.02) | 0.1 (0.06) | 0.66 (0.02) | 0.13 (0.07) | 0.6 (0.02) | 0.24 (0.06) | 0.15 (0.03) | 0.1 (0.06) | 0.18 (0.04) | 0.16 (0.07) | 0.16 (0.04) | 0.07 (0.07) | 0.2 (0.04) | 0.09 (0.08) | 0.18 (0.04) | 0.2 (0.08) | 0.18 (0.03) | 0.06 (0.07) |
| | Cig T3 | 0.17 (0.07) | 0.76 (0.05) | 0.11 (0.08) | 0.92 (0.02) | 0.13 (0.07) | 1 | 0.24 (0.06) | 0.88 (0.03) | 0.27 (0.06) | 0.92 (0.02) | 0.2 (0.05) | 0.84 (0.03) | 0.08 (0.06) | 0.39 (0.08) | 0.15 (0.07) | 0.41 (0.1) | 0.03 (0.08) | 0.45 (0.1) | 0.16 (0.08) | 0.36 (0.12) | 0.12 (0.08) | 0.52 (0.1) | 0.04 (0.06) | 0.32 (0.1) |
| | Dep T4 | 0.58 (0.03) | 0.01 (0.08) | 0.64 (0.02) | -0.13 (0.08) | 0.7 (0.02) | -0.07 (0.09) | 1 | 0.11 (0.06) | 0.7 (0.02) | 0.18 (0.07) | 0.62 (0.02) | 0.3 (0.06) | 0.21 (0.03) | 0.12 (0.06) | 0.21 (0.04) | 0.08 (0.08) | 0.24 (0.04) | 0.06 (0.08) | 0.19 (0.04) | 0.16 (0.08) | 0.25 (0.04) | 0.12 (0.09) | 0.19 (0.03) | 0.02 (0.07) |
| | Cig T4 | 0.21 (0.08) | 0.69 (0.06) | 0.08 (0.08) | 0.82 (0.04) | 0.07 (0.09) | 0.84 (0.04) | 0.07 (0.07) | 1 | 0.13 (0.06) | 0.88 (0.02) | 0.08 (0.06) | 0.79 (0.04) | 0.07 (0.06) | 0.35 (0.09) | 0.04 (0.08) | 0.38 (0.11) | 0.06 (0.09) | 0.3 (0.12) | 0.09 (0.08) | 0.33 (0.12) | 0.11 (0.08) | 0.45 (0.11) | 0.03 (0.07) | 0.24 (0.11) |
| | Dep T5 | 0.5 (0.03) | 0.04 (0.08) | 0.65 (0.02) | 0.07 (0.08) | 0.64 (0.03) | 0.11 (0.08) | 0.68 (0.02) | 0.14 (0.09) | 1 | 0.25 (0.05) | 0.64 (0.02) | 0.32 (0.06) | 0.2 (0.03) | 0.14 (0.06) | 0.18 (0.04) | 0 (0.07) | 0.19 (0.04) | 0.06 (0.07) | 0.18 (0.04) | 0.11 (0.08) | 0.23 (0.04) | 0.1 (0.08) | 0.18 (0.03) | 0.03 (0.07) |
| | Cig T5 | 0.16 (0.08) | 0.7 (0.07) | 0 (0.09) | 0.88 (0.03) | -0.01 (0.09) | 0.85 (0.04) | -0.13 (0.09) | 0.93 (0.02) | 0.05 (0.08) | 1 | 0.16 (0.06) | 0.87 (0.02) | 0.02 (0.06) | 0.36 (0.08) | 0.07 (0.08) | 0.3 (0.12) | 0.1 (0.08) | 0.36 (0.12) | 0.16 (0.08) | 0.4 (0.12) | 0.03 (0.08) | 0.52 (0.1) | -0.05 (0.06) | 0.38 (0.1) |

| | | Twin 1 | | | | | | | | | | | | Twin 2 | | | | | | | | | | | |
|---|---|---|---|---|---|---|---|---|---|---|---|---|---|---|---|---|---|---|---|---|---|---|---|---|---|
| | | **DepT1** | **CigT1** | **DepT2** | **CigT2** | **DepT3** | **CigT3** | **DepT4** | **CigT4** | **DepT5** | **CigT5** | **DepT6** | **CigT6** | **DepT1** | **CigT1** | **DepT2** | **CigT2** | **DepT3** | **CigT3** | **DepT4** | **CigT4** | **DepT5** | **CigT5** | **DepT6** | **CigT6** |
| | **Dep T6** | 0.55 (0.02) | 0.14 (0.06) | 0.56 (0.02) | 0.1 (0.07) | 0.58 (0.03) | 0.15 (0.08) | 0.6 (0.03) | 0.21 (0.08) | 0.64 (0.02) | 0.17 (0.08) | 1 | 0.23 (0.04) | 0.17 (0.02) | 0.12 (0.04) | 0.2 (0.03) | 0.11 (0.06) | 0.16 (0.03) | 0.11 (0.06) | 0.16 (0.04) | 0.15 (0.07) | 0.2 (0.03) | 0.1 (0.07) | 0.24 (0.02) | 0.04 (0.05) |
| | **Cig T6** | 0.04 (0.06) | 0.7 (0.05) | 0.13 (0.08) | 0.83 (0.04) | 0.12 (0.08) | 0.81 (0.04) | -0.07 (0.08) | 0.85 (0.04) | 0.1 (0.09) | 0.92 (0.02) | 0.22 (0.05) | 1 | 0.02 (0.05) | 0.27 (0.07) | 0.12 (0.07) | 0.23 (0.1) | 0.14 (0.07) | 0.22 (0.11) | 0.13 (0.08) | 0.14 (0.12) | 0.1 (0.07) | 0.35 (0.11) | 0.03 (0.05) | 0.31 (0.08) |
| **Twin 2** | **Dep T1** | 0.38 (0.02) | 0.17 (0.05) | 0.37 (0.03) | 0.01 (0.07) | 0.36 (0.03) | 0.03 (0.08) | 0.33 (0.04) | 0.05 (0.08) | 0.31 (0.04) | 0 (0.09) | 0.33 (0.03) | 0.09 (0.06) | 1 | 0.21 (0.03) | 0.5 (0.02) | 0.2 (0.05) | 0.55 (0.02) | 0.2 (0.06) | 0.54 (0.02) | 0.19 (0.06) | 0.51 (0.03) | 0.13 (0.06) | 0.51 (0.02) | 0.2 (0.04) |
| | **Cig T1** | 0.07 (0.05) | 0.65 (0.05) | 0.1 (0.07) | 0.61 (0.07) | 0.01 (0.08) | 0.54 (0.09) | -0.01 (0.08) | 0.52 (0.09) | 0.09 (0.08) | 0.54 (0.1) | 0.07 (0.06) | 0.64 (0.06) | 0.23 (0.05) | 1 | 0.21 (0.05) | 0.73 (0.04) | 0.18 (0.06) | 0.76 (0.04) | 0.25 (0.06) | 0.71 (0.05) | 0.26 (0.06) | 0.73 (0.05) | 0.26 (0.04) | 0.76 (0.03) |
| | **Dep T2** | 0.4 (0.03) | 0.03 (0.07) | 0.41 (0.03) | 0.07 (0.08) | 0.47 (0.03) | 0.02 (0.09) | 0.43 (0.04) | 0.07 (0.09) | 0.4 (0.04) | 0.06 (0.09) | 0.39 (0.03) | 0.17 (0.08) | 0.58 (0.02) | 0.13 (0.07) | 1 | 0.2 (0.05) | 0.69 (0.02) | 0.2 (0.06) | 0.63 (0.02) | 0.26 (0.06) | 0.63 (0.02) | 0.24 (0.07) | 0.57 (0.02) | 0.16 (0.07) |
| | **Cig T2** | 0.12 (0.07) | 0.55 (0.08) | 0.06 (0.08) | 0.6 (0.08) | 0.08 (0.09) | 0.46 (0.11) | -0.05 (0.09) | 0.54 (0.1) | 0.08 (0.09) | 0.53 (0.1) | 0.11 (0.07) | 0.61 (0.08) | 0.1 (0.07) | 0.82 (0.04) | 0.17 (0.06) | 1 | 0.13 (0.07) | 0.9 (0.02) | 0.19 (0.07) | 0.81 (0.04) | 0.3 (0.07) | 0.8 (0.04) | 0.24 (0.06) | 0.77 (0.04) |
| | **Dep T3** | 0.39 (0.03) | -0.06 (0.08) | 0.38 (0.04) | 0.04 (0.09) | 0.39 (0.04) | 0.01 (0.09) | 0.36 (0.04) | -0.03 (0.1) | 0.34 (0.04) | 0.04 (0.1) | 0.34 (0.04) | 0.17 (0.08) | 0.61 (0.02) | 0.19 (0.07) | 0.75 (0.02) | 0.14 (0.08) | 1 | 0.1 (0.06) | 0.71 (0.02) | 0.24 (0.07) | 0.68 (0.02) | 0.11 (0.08) | 0.63 (0.02) | 0.14 (0.07) |
| | **Cig T3** | 0.1 (0.07) | 0.61 (0.08) | 0.05 (0.08) | 0.65 (0.07) | 0.03 (0.09) | 0.44 (0.11) | -0.06 (0.09) | 0.57 (0.1) | 0 (0.09) | 0.57 (0.1) | 0.05 (0.08) | 0.61 (0.08) | 0.1 (0.07) | 0.77 (0.05) | 0.16 (0.07) | 0.93 (0.02) | 0.16 (0.07) | 1 | 0.11 (0.07) | 0.86 (0.03) | 0.24 (0.07) | 0.83 (0.04) | 0.2 (0.06) | 0.83 (0.04) |
| | **Dep T4** | 0.39 (0.04) | 0.02 (0.08) | 0.41 (0.04) | 0.05 (0.1) | 0.4 (0.04) | 0 (0.1) | 0.37 (0.04) | -0.02 (0.1) | 0.4 (0.04) | -0.04 (0.11) | 0.38 (0.04) | 0.01 (0.09) | 0.58 (0.03) | 0.15 (0.08) | 0.66 (0.02) | 0.04 (0.09) | 0.71 (0.02) | 0.11 (0.09) | 1 | 0.25 (0.05) | 0.68 (0.02) | 0.2 (0.07) | 0.63 (0.02) | 0.19 (0.07) |
| | **Cig T4** | 0.11 (0.08) | 0.57 (0.09) | 0.08 (0.09) | 0.53 (0.11) | 0.1 (0.09) | 0.47 (0.11) | 0.06 (0.09) | 0.65 (0.09) | 0.17 (0.09) | 0.55 (0.11) | 0.2 (0.08) | 0.59 (0.09) | 0.14 (0.08) | 0.7 (0.06) | 0.17 (0.08) | 0.86 (0.03) | 0.08 (0.09) | 0.86 (0.03) | 0.09 (0.07) | 1 | 0.31 (0.07) | 0.85 (0.03) | 0.29 (0.06) | 0.8 (0.03) |
| | **Dep T5** | 0.39 (0.03) | 0.07 (0.08) | 0.39 (0.04) | 0.12 (0.09) | 0.35 (0.04) | 0.03 (0.09) | 0.37 (0.04) | 0.08 (0.09) | 0.36 (0.04) | 0.02 (0.1) | 0.4 (0.03) | 0.11 (0.09) | 0.61 (0.02) | 0.14 (0.08) | 0.64 (0.02) | 0.19 (0.08) | 0.68 (0.02) | 0.21 (0.08) | 0.71 (0.02) | 0.06 (0.09) | 1 | 0.23 (0.06) | 0.67 (0.02) | 0.18 (0.07) |

| | Twin 1 | | | | | | | | | | | | Twin 2 | | | | | | | | | | | |
|---|---|---|---|---|---|---|---|---|---|---|---|---|---|---|---|---|---|---|---|---|---|---|---|---|
| | **DepT1** | **CigT1** | **DepT2** | **CigT2** | **DepT3** | **CigT3** | **DepT4** | **CigT4** | **DepT5** | **CigT5** | **DepT6** | **CigT6** | **DepT1** | **CigT1** | **DepT2** | **CigT2** | **DepT3** | **CigT3** | **DepT4** | **CigT4** | **DepT5** | **CigT5** | **DepT6** | **CigT6** |
| **Cig T5** | 0.12 (0.08) | 0.67 (0.07) | -0.02 (0.09) | 0.67 (0.08) | 0.1 (0.1) | 0.61 (0.09) | 0.06 (0.1) | 0.71 (0.07) | 0.12 (0.09) | 0.62 (0.1) | 0.21 (0.08) | 0.65 (0.08) | 0.2 (0.08) | 0.7 (0.06) | 0.19 (0.08) | 0.87 (0.04) | 0.15 (0.08) | 0.84 (0.04) | 0.04 (0.09) | 0.89 (0.02) | 0.25 (0.07) | 1 | 0.15 (0.06) | 0.85 (0.03) |
| **Dep T6** | 0.4 (0.03) | 0.23 (0.06) | 0.37 (0.03) | 0.13 (0.07) | 0.35 (0.04) | 0.07 (0.08) | 0.34 (0.04) | 0.07 (0.08) | 0.31 (0.04) | 0.13 (0.08) | 0.41 (0.03) | 0.19 (0.06) | 0.53 (0.02) | 0.2 (0.05) | 0.56 (0.03) | 0.2 (0.07) | 0.64 (0.02) | 0.18 (0.07) | 0.65 (0.02) | 0.11 (0.08) | 0.68 (0.02) | 0.21 (0.07) | 1 | 0.24 (0.04) |
| **Cig T6** | 0.12 (0.06) | 0.65 (0.06) | 0.11 (0.08) | 0.72 (0.07) | 0.09 (0.09) | 0.69 (0.08) | 0.02 (0.09) | 0.78 (0.06) | 0.08 (0.09) | 0.72 (0.07) | 0.21 (0.06) | 0.76 (0.05) | 0.23 (0.06) | 0.75 (0.04) | 0.21 (0.07) | 0.84 (0.04) | 0.23 (0.08) | 0.84 (0.04) | 0.1 (0.09) | 0.9 (0.02) | 0.2 (0.08) | 0.94 (0.02) | 0.19 (0.05) | 1 |

*Note.* *The bottom off-diagonal triangle (in blue) shows the correlations in MZ twins, while the top triangle (in yellow) shows the correlations in DZ twins. The values in parentheses indicate the standard error of the estimated correlation. Dep = DepSx. Cig = CigDay. T1–T6 indicate TEDS-21, COVID Phase 1–4, and TEDS-26, respectively.*

**Table S4**

*Parameter Estimates in Standard Twin CLPM Applied to TEDS-21 and TEDS-26*

| | Parameter | Description | Estimate | Std.Error |
|---|---|---|---|---|
| 1 | *b0_DepSx_T1* | Mean DepSx TEDS-21 | 0.027 | 0.013 |
| 2 | *b0_CigDay_T1* | Mean CigDay TEDS-21 | -0.012 | 0.029 |
| 3 | *b0_DepSx_T2* | Mean DepSx TEDS-26 | -0.040 | 0.014 |
| 4 | *b0_CigDay_T2* | Mean CigDay TEDS-26 | -0.122 | 0.035 |
| 5 | *VAx1* | VA of DepSx at TEDS-21 | 0.282 | 0.016 |
| 6 | *covA1* | covA at TEDS-21 | 0.155 | 0.018 |
| 7 | *VAy1* | VA of CigDay at TEDS-21 | 0.371 | 0.035 |
| 8 | *VAx2* | VA of DepSx at TEDS-26 | 0.193 | 0.019 |
| 9 | *covA2* | covA at TEDS-26 | 0.020 | 0.025 |
| 10 | *VAy2* | VA of CigDay at TEDS-26 | 0.130 | 0.023 |
| 11 | *VEx1* | VE of DepSx at TEDS-21 | 0.477 | 0.014 |
| 12 | *covE1* | covE at TEDS-21 | -0.010 | 0.015 |
| 13 | *VEy1* | VE of CigDay at TEDS-21 | 0.118 | 0.018 |
| 14 | *VEx2* | VE of DepSx at TEDS-26 | 0.580 | 0.018 |
| 15 | *covE2* | covE at TEDS-26 | 0.030 | 0.026 |
| 16 | *VEy2* | VE of CigDay at TEDS-26 | 0.071 | 0.020 |
| 17 | *arX* | AR of DepSx | 0.552 | 0.013 |
| 18 | ***by2x1*** | DepSx → CigDay | **-0.003** | **0.020** |
| 19 | ***bx2y1*** | CigDay → DepSx | **0.223** | **0.032** |
| 20 | *arY* | AR of CigDay | 0.764 | 0.044 |

**Table S5**

*Parameter Estimates in the Best-Fitting Standard Twin CLPM Applied to TEDS-21 and TEDS-26*

| | Parameter | Description | Estimate | Std.Error |
|---|---|---|---|---|
| 1 | *b0_DepSx_T1* | Mean DepSx TEDS-21 | 0.027 | 0.013 |
| 2 | *b0_CigDay_T1* | Mean CigDay TEDS-21 | -0.013 | 0.029 |
| 3 | *b0_DepSx_T2* | Mean DepSx TEDS-26 | -0.040 | 0.014 |
| 4 | *b0_CigDay_T2* | Mean CigDay TEDS-26 | -0.122 | 0.035 |
| 5 | *VAx1* | VA of DepSx at TEDS-21 | 0.282 | 0.016 |
| 6 | *covA1* | covA at TEDS-21 | 0.146 | 0.013 |
| 7 | *VAy1* | VA of CigDay at TEDS-21 | 0.371 | 0.035 |
| 8 | *VAx2* | VA of DepSx at TEDS-26 | 0.193 | 0.019 |
| 9 | *VAy2* | VA of CigDay at TEDS-26 | 0.127 | 0.023 |
| 10 | *VEx1* | VE of DepSx at TEDS-21 | 0.477 | 0.014 |
| 11 | *VEy1* | VE of CigDay at TEDS-21 | 0.119 | 0.018 |
| 12 | *VEx2* | VE of DepSx at TEDS-26 | 0.580 | 0.018 |
| 13 | *covE2* | covE at TEDS-26 | 0.047 | 0.015 |
| 14 | *VEy2* | VE of CigDay at TEDS-26 | 0.073 | 0.020 |
| 15 | *arX* | AR of DepSx | 0.552 | 0.012 |
| 16 | ***bx2y1*** | CigDay → DepSx | **0.220** | **0.032** |
| 17 | *arY* | AR of CigDay | 0.763 | 0.044 |

**Table S6**

*Parameter Estimates in the Biometrical CLPM Applied to TEDS-21 and TEDS-26*

| | Parameter | Description | Estimate | Std.Error |
|---|---|---|---|---|
| 1 | *b0_DepSx_T1* | Mean DepSx TEDS-21 | 0.025 | 0.013 |
| 2 | *b0_CigDay_T1* | Mean CigDay TEDS-21 | -0.001 | 0.027 |
| 3 | *b0_DepSx_T2* | Mean DepSx TEDS-26 | -0.035 | 0.014 |
| 4 | *b0_CigDay_T2* | Mean CigDay TEDS-26 | -0.121 | 0.035 |
| 5 | *VAx1* | VA of DepSx at TEDS-21 | 0.277 | 0.016 |
| 6 | *covA1* | covA at TEDS-21 | 0.080 | 0.018 |
| 7 | *VAy1* | VA of CigDay at TEDS-21 | 0.311 | 0.032 |
| 8 | *VAx2* | VA of DepSx at TEDS-26 | 0.081 | 0.026 |
| 9 | *covA2* | covA at TEDS-26 | 0.016 | 0.026 |
| 10 | *VAy2* | VA of CigDay at TEDS-26 | 0.062 | 0.032 |
| 11 | arAx | AR of A_DepSx | 1.141 | 0.060 |
| 12 | arAy | AR of A_CigDay | 0.975 | 0.086 |
| 13 | *VEx1* | VE of DepSx at TEDS-21 | 0.502 | 0.014 |
| 14 | *covE1* | covE at TEDS-21 | 0.046 | 0.016 |
| 15 | *VEy1* | VE of CigDay at TEDS-21 | 0.145 | 0.019 |
| 16 | *VEx2* | VE of DepSx at TEDS-26 | 0.601 | 0.018 |
| 17 | *covE2* | covE at TEDS-26 | 0.037 | 0.025 |
| 18 | *VEy2* | VE of CigDay at TEDS-26 | 0.103 | 0.020 |
| 19 | arEx | AR of E_DepSx | 0.313 | 0.024 |
| 20 | arEy | AR of E_CigDay | 0.330 | 0.117 |
| 21 | ***by2x1*** | DepSx → CigDay | **0.004** | **0.022** |
| 22 | ***bx2y1*** | CigDay → DepSx | **0.069** | **0.044** |

**Table S7**

*Parameter Estimates in the Best-Fitting Biometrical CLPM with Unidirectional Proximal Causation*

| | Parameter | Description | Estimate | Std.Error |
|---|---|---|---|---|
| 1 | *b0_DepSx_T1* | Mean DepSx TEDS-21 | 0.025 | 0.013 |
| 2 | *b0_CigDay_T1* | Mean CigDay TEDS-21 | -0.001 | 0.027 |
| 3 | *b0_DepSx_T2* | Mean DepSx TEDS-26 | -0.035 | 0.014 |
| 4 | *b0_CigDay_T2* | Mean CigDay TEDS-26 | -0.121 | 0.035 |
| 5 | *VAx1* | VA of DepSx at TEDS-21 | 0.277 | 0.016 |
| 6 | *covA1* | covA at TEDS-21 | 0.080 | 0.015 |
| 7 | *VAy1* | VA of CigDay at TEDS-21 | 0.311 | 0.032 |
| 8 | *VAx2* | VA of DepSx at TEDS-26 | 0.081 | 0.026 |
| 9 | *VAy2* | VA of CigDay at TEDS-26 | 0.062 | 0.032 |
| 10 | arAx | AR of A_DepSx | 1.141 | 0.061 |
| 11 | arAy | AR of A_CigDay | 0.975 | 0.086 |
| 12 | *VEx1* | VE of DepSx at TEDS-21 | 0.502 | 0.014 |
| 13 | *covE1* | covE at TEDS-21 | 0.046 | 0.014 |
| 14 | *VEy1* | VE of CigDay at TEDS-21 | 0.145 | 0.019 |
| 15 | *VEx2* | VE of DepSx at TEDS-26 | 0.601 | 0.019 |
| 16 | *covE2* | covE at TEDS-26 | 0.037 | 0.019 |
| 17 | *VEy2* | VE of CigDay at TEDS-26 | 0.103 | 0.020 |
| 18 | arEx | AR of E_DepSx | 0.313 | 0.024 |
| 19 | arEy | AR of E_CigDay | 0.330 | 0.115 |
| 20 | ***bx2y2*** | **Proximal CigDay → DepSx** | **0.100** | **0.050** |

**Table S8**

*Parameter Estimates in the Biometrical CLPM with Bidirectional Proximal Causation*

| | Parameter | Description | Estimate | Std.Error |
|---|---|---|---|---|
| 1 | *b0_DepSx_T1* | Mean DepSx TEDS-21 | 0.024 | 0.013 |
| 2 | *b0_CigDay_T1* | Mean CigDay TEDS-21 | 0.000 | 0.027 |
| 3 | *b0_DepSx_T2* | Mean DepSx TEDS-26 | -0.035 | 0.014 |
| 4 | *b0_CigDay_T2* | Mean CigDay TEDS-26 | -0.119 | 0.035 |
| 5 | *VAx1* | VA of DepSx at TEDS-21 | 0.276 | 0.016 |
| 6 | *covA1* | covA at TEDS-21 | 0.072 | 0.015 |
| 7 | *VAy1* | VA of CigDay at TEDS-21 | 0.309 | 0.032 |
| 8 | *VAx2* | VA of DepSx at TEDS-26 | 0.077 | 0.026 |
| 9 | *VAy2* | VA of CigDay at TEDS-26 | 0.056 | 0.031 |
| 10 | arAx | AR of A_DepSx | 1.130 | 0.060 |
| 11 | arAy | AR of A_CigDay | 0.976 | 0.086 |
| 12 | *VEx1* | VE of DepSx at TEDS-21 | 0.503 | 0.014 |
| 13 | *covE1* | covE at TEDS-21 | 0.052 | 0.014 |
| 14 | *VEy1* | VE of CigDay at TEDS-21 | 0.146 | 0.019 |
| 15 | *VEx6* | VE of DepSx at TEDS-26 | 0.596 | 0.018 |
| 16 | *VEy6* | VE of CigDay at TEDS-26 | 0.103 | 0.019 |
| 17 | arEx | AR of E_DepSx | 0.315 | 0.023 |
| 18 | arEy | AR of E_CigDay | 0.308 | 0.112 |
| 19 | ***by2x2*** | **Proximal DepSx → CigDay** | **0.029** | **0.018** |
| 20 | ***bx2y2*** | **Proximal CigDay → DepSx** | **0.119** | **0.043** |

**Table S9**

*Model Selection in the Biometrical CLPM with Six Study Waves*

| Base Model | Comparison Model | EP | Browne's Pseudo-$\chi^2$ | DF | AIC | ΔAIC | SB $\chi^2$ | ΔDF | SB $\chi^2$ p-value |
|---|---|---|---|---|---|---|---|---|---|
| **AE autoregression; Bidirectional cross-lagged effects** | | **78** | **2673.03** | **570** | **2829.03** | | | | |
| AE autoregression; Bidirectional cross-lagged effects | Phenotypic autoregression | 68 | 3640.97 | 580 | 3776.97 | 947.94 | 429.21 | 10 | <0.0001 |
| AE autoregression; Bidirectional cross-lagged effects | No cross-lagged associations | 68 | 2874.04 | 580 | 3010.04 | 181.00 | 82.69 | 10 | <0.0001 |
| AE autoregression; Bidirectional cross-lagged effects | Unidirectional lagged *DepSx* → *CigDay* effects | 73 | 2799.77 | 575 | 2945.77 | 116.74 | 52.29 | 5 | <0.0001 |
| AE autoregression; Bidirectional cross-lagged effects | Unidirectional lagged *CigDay* → *DepSx* effects | 73 | 2861.09 | 575 | 3007.09 | 178.06 | 75.87 | 5 | <0.0001 |

*Note. EP = The number of estimated parameters. DF = Degrees of freedom. AIC = Akaike information criterion. SB $\chi^2$ = Satorra-Bentler scaled-difference $\chi^2$ statistic.*

**Table S10**

*Parameter Estimates in the Six-Wave Biometrical CLPM Applied to TEDS-21, COVID Study Phases 1–4, and TEDS-26*

| | Parameter | Description | Estimate | Std.Error |
|---|---|---|---|---|
| 1 | *b0_DepSx_T1* | Mean DepSx TEDS-21 | 0.067 | 0.012 |
| 2 | *b0_CigDay_T1* | Mean CigDay TEDS-21 | -0.030 | 0.025 |
| 3 | *b0_DepSx_T2* | Mean DepSx COVID-1 | 0.082 | 0.014 |
| 4 | *b0_CigDay_T2* | Mean CigDay COVID-1 | 0.094 | 0.026 |
| 5 | *b0_DepSx_T3* | Mean DepSx COVID-2 | 0.039 | 0.016 |
| 6 | *b0_CigDay_T3* | Mean CigDay COVID-2 | 0.162 | 0.022 |
| 7 | *b0_DepSx_T4* | Mean DepSx COVID-3 | 0.040 | 0.017 |
| 8 | *b0_CigDay_T4* | Mean CigDay COVID-3 | 0.170 | 0.023 |
| 9 | *b0_DepSx_T5* | Mean DepSx COVID-4 | -0.049 | 0.017 |
| 10 | *b0_CigDay_T5* | Mean CigDay COVID-4 | -0.055 | 0.037 |
| 11 | *b0_DepSx_T6* | Mean DepSx TEDS-26 | -0.053 | 0.013 |
| 12 | *b0_CigDay_T6* | Mean CigDay TEDS-26 | -0.237 | 0.039 |
| 13 | *VAx1* | VA of DepSx at TEDS-21 | 0.228 | 0.013 |
| 14 | *covA1* | covA at TEDS-21 | -0.069 | 0.008 |
| 15 | *VAy1* | VA of CigDay at TEDS-21 | 0.323 | 0.025 |
| 16 | *VAx2* | VA of DepSx at COVID-1 | -0.006 | 0.015 |
| 17 | *covA2* | covA at COVID-1 | -0.086 | 0.008 |
| 18 | *VAy2* | VA of CigDay at COVID-1 | 0.049 | 0.008 |
| 19 | *VAx3* | VA of DepSx at COVID-2 | 0.005 | 0.010 |
| 20 | *covA3* | covA at COVID-2 | -0.032 | 0.006 |
| 21 | *VAy3* | VA of CigDay at COVID-2 | 0.013 | 0.005 |
| 22 | *VAx4* | VA of DepSx at COVID-3 | -0.005 | 0.009 |
| 23 | *covA4* | covA at COVID-3 | 0.004 | 0.005 |
| 24 | *VAy4* | VA of CigDay at COVID-3 | 0.018 | 0.004 |
| 25 | *VAx5* | VA of DepSx at COVID-4 | 0.006 | 0.011 |
| 26 | *covA5* | covA at COVID-4 | -0.016 | 0.011 |

| | Parameter | Description | Estimate | Std.Error |
|---|---|---|---|---|
| 27 | *VAy5* | VA of CigDay at COVID-4 | -0.049 | 0.013 |
| 28 | *VAx6* | VA of DepSx at TEDS-26 | 0.062 | 0.019 |
| 29 | *covA6* | covA at TEDS-26 | 0.056 | 0.024 |
| 30 | *VAy6* | VA of CigDay at TEDS-26 | -0.009 | 0.024 |
| 31 | *AR_Ax2x1* | AR of A_DepSex T1 → T2 | 1.241 | 0.063 |
| 32 | *AR_Ay2y1* | AR of A_CigDay T1 → T2 | 0.818 | 0.045 |
| 33 | *AR_Ax3x2* | AR of A_DepSex T2 → T3 | 1.194 | 0.032 |
| 34 | *AR_Ay3y2* | AR of A_CigDay T2 → T3 | 0.967 | 0.046 |
| 35 | *AR_Ax4x3* | AR of A_DepSex T3 → T4 | 1.083 | 0.023 |
| 36 | *AR_Ay4y3* | AR of A_CigDay T3 → T4 | 0.735 | 0.038 |
| 37 | *AR_Ax5x4* | AR of A_DepSex T4 → T5 | 1.028 | 0.021 |
| 38 | *AR_Ay5y4* | AR of A_CigDay T4 → T5 | 1.632 | 0.093 |
| 39 | *AR_Ax6x5* | AR of A_DepSex T5 → T6 | 0.933 | 0.022 |
| 40 | *AR_Ay6y5* | AR of A_CigDay T5 → T6 | 1.150 | 0.069 |
| 41 | *VEx1* | VE of DepSx at TEDS-21 | 0.439 | 0.011 |
| 42 | *covE1* | covE at TEDS-21 | 0.145 | 0.010 |
| 43 | *VEy1* | VE of CigDay at TEDS-21 | 0.141 | 0.014 |
| 44 | *VEx2* | VE of DepSx at COVID-1 | 0.379 | 0.010 |
| 45 | *covE2* | covE at COVID-1 | 0.099 | 0.008 |
| 46 | *VEy2* | VE of CigDay at COVID-1 | 0.067 | 0.007 |
| 47 | *VEx3* | VE of DepSx at COVID-2 | 0.287 | 0.008 |
| 48 | *covE3* | covE at COVID-2 | 0.040 | 0.006 |
| 49 | *VEy3* | VE of CigDay at COVID-2 | 0.007 | 0.004 |
| 50 | *VEx4* | VE of DepSx at COVID-3 | 0.348 | 0.009 |
| 51 | *covE4* | covE at COVID-3 | 0.027 | 0.008 |
| 52 | *VEy4* | VE of CigDay at COVID-3 | -0.005 | 0.010 |
| 53 | *VEx5* | VE of DepSx at COVID-4 | 0.334 | 0.013 |
| 54 | *covE5* | covE at COVID-4 | 0.021 | 0.015 |
| 55 | *VEy5* | VE of CigDay at COVID-4 | 0.067 | 0.010 |
| 56 | *VEx6* | VE of DepSx at TEDS-26 | 0.406 | 0.016 |

| | Parameter | Description | Estimate | Std.Error |
|---|---|---|---|---|
| 57 | *covE6* | covE at TEDS-26 | -0.021 | 0.023 |
| 58 | *VEy6* | VE of CigDay at TEDS-26 | 0.042 | 0.021 |
| 59 | *AR_Ex2x1* | AR of E_DepSex T1 → T2 | 0.116 | 0.021 |
| 60 | *AR_Ey2y1* | AR of E_CigDay T1 → T2 | 0.516 | 0.051 |
| 61 | *AR_Ex3x2* | AR of E_DepSex T2 → T3 | 0.258 | 0.021 |
| 62 | *AR_Ey3y2* | AR of E_CigDay T2 → T3 | 0.695 | 0.066 |
| 63 | *AR_Ex4x3* | AR of E_DepSex T3 → T4 | 0.210 | 0.019 |
| 64 | *AR_Ey4y3* | AR of E_CigDay T3 → T4 | 1.567 | 0.134 |
| 65 | *AR_Ex5x4* | AR of E_DepSex T4 → T5 | 0.192 | 0.027 |
| 66 | *AR_Ey5y4* | AR of E_CigDay T4 → T5 | 0.850 | 0.053 |
| 67 | *AR_Ex6x5* | AR of E_DepSex T5 → T6 | 0.184 | 0.036 |
| 68 | *AR_Ey6y5* | AR of E_CigDay T5 → T6 | 0.609 | 0.062 |
| 69 | ***by2x1*** | **DepSx_T1 → CigDay_T2** | **0.063** | **0.018** |
| 70 | ***bx2y1*** | **CigDay_T1 → DepSx_T2** | **0.339** | **0.034** |
| 71 | ***by3x2*** | **DepSx_T2 → CigDay_T3** | **0.106** | **0.016** |
| 72 | ***bx3y2*** | **CigDay_T2 → DepSx_T3** | **0.608** | **0.044** |
| 73 | ***by4x3*** | **DepSx_T3 → CigDay_T4** | **-0.040** | **0.016** |
| 74 | ***bx4y3*** | **CigDay_T3 → DepSx_T4** | **0.724** | **0.046** |
| 75 | ***by5x4*** | **DepSx_T4 → CigDay_T5** | **0.114** | **0.018** |
| 76 | ***bx5y4*** | **CigDay_T4 → DepSx_T5** | **0.996** | **0.058** |
| 77 | ***by6x5*** | **DepSx_T5 → CigDay_T6** | **0.178** | **0.024** |
| 78 | ***bx6y5*** | **CigDay_T5 → DepSx_T6** | **0.704** | **0.045** |

### Supplementary Figures

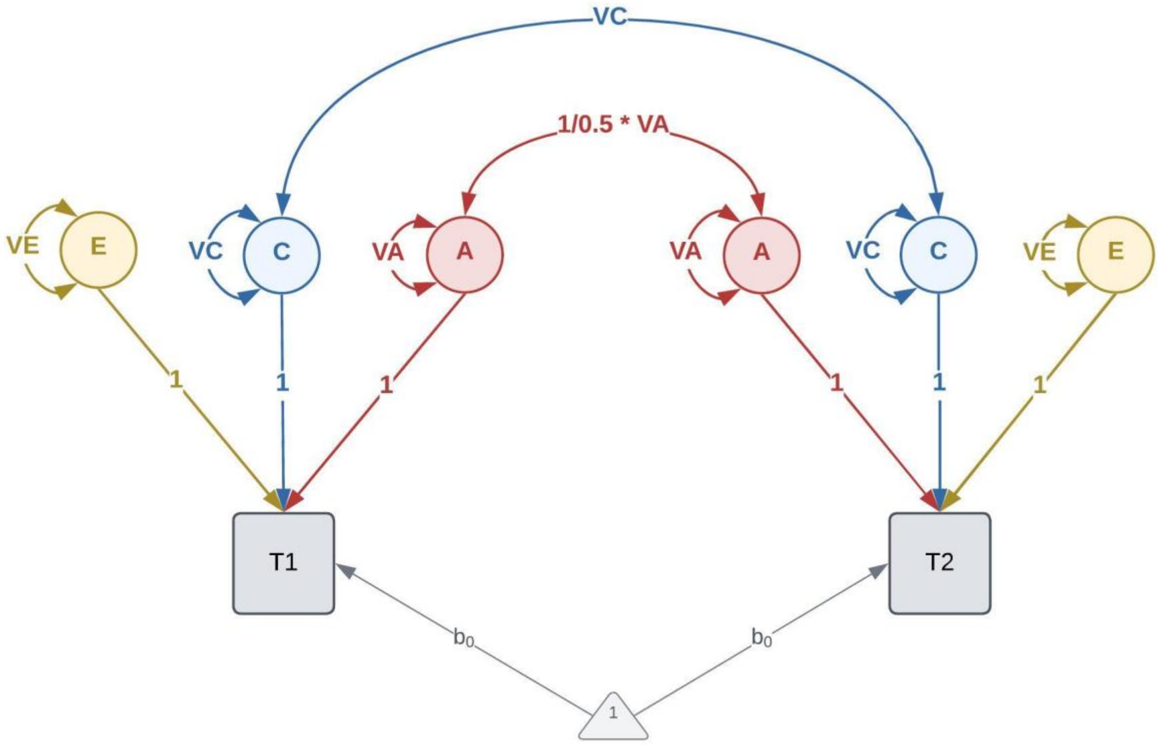

**Figure S1. Univariate Twin Model**

*The variance of a phenotype observed in pairs of twins (T1 and T2) is decomposed into latent additive genetic (A), shared familial environmental (C), and unique individual-specific environmental (E) factors. The twin correlation of the additive genetic factors is 1 in MZ and 0.5 in DZ twin pairs. The twin correlation of the familial environmental factors is 1 in both MZ and DZ pairs. The squares/rectangles indicate observed variables, the circles indicate latent (unobserved) variables, the single-headed arrows indicate regression paths, and the double-headed curved arrows indicate (co-)variances. The triangle indicates a constant value of 1, and the paths from this constant to the observed variables are used to estimate the mean ($b_0$).*

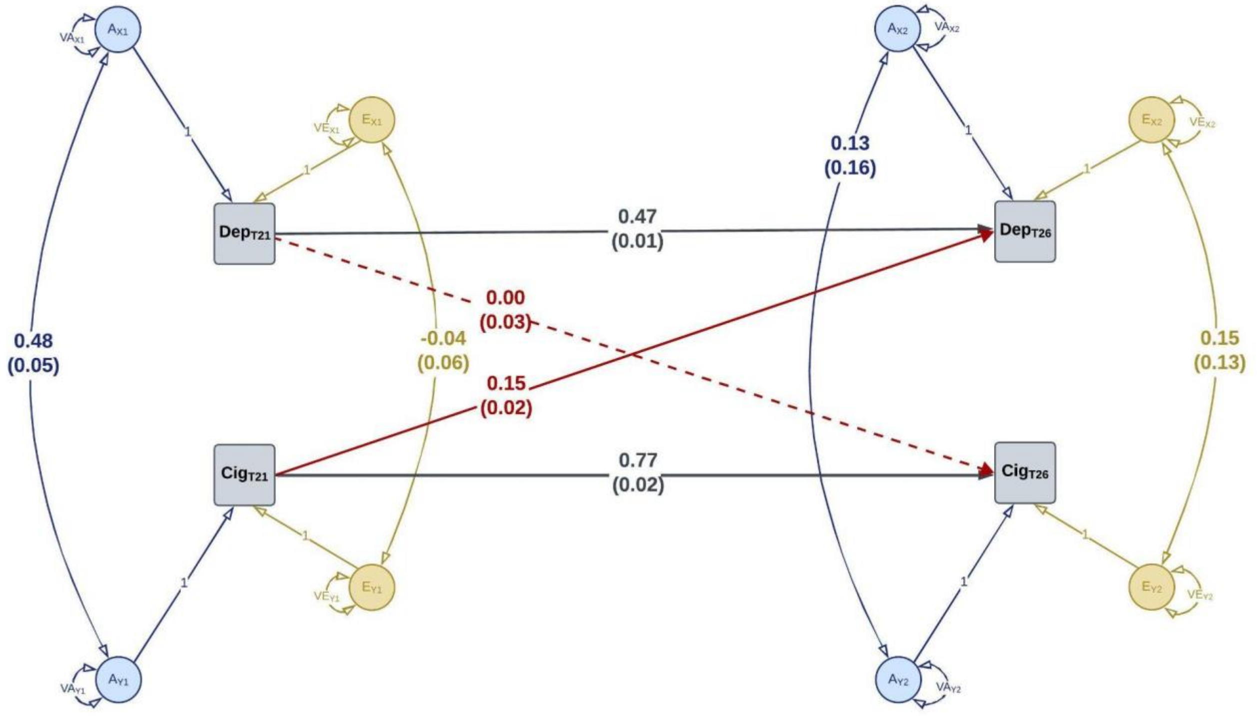

**Figure S2. Standard twin CLPM (Full Model).**

*Note. Standardized estimates of selected parameters are shown. The dashed line indicates a non-significant estimate. All parameter estimates are shown in* ***Supplementary Table S4****.*

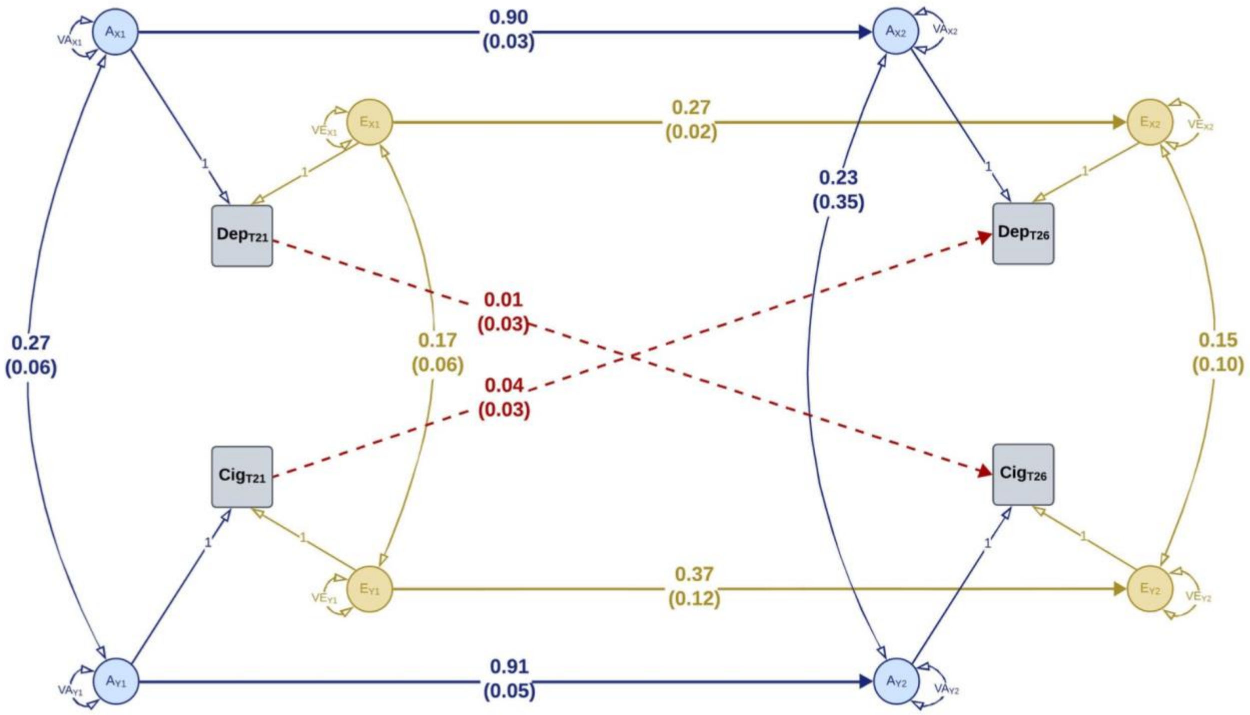

**Figure S3. Biometrical CLPM (Full Model with Cross-Lagged Causation).**

*Note. Standardized estimates of selected parameters are shown. The dashed line indicates a non-significant estimate. All parameter estimates are shown in* ***Supplementary Table S6****.*

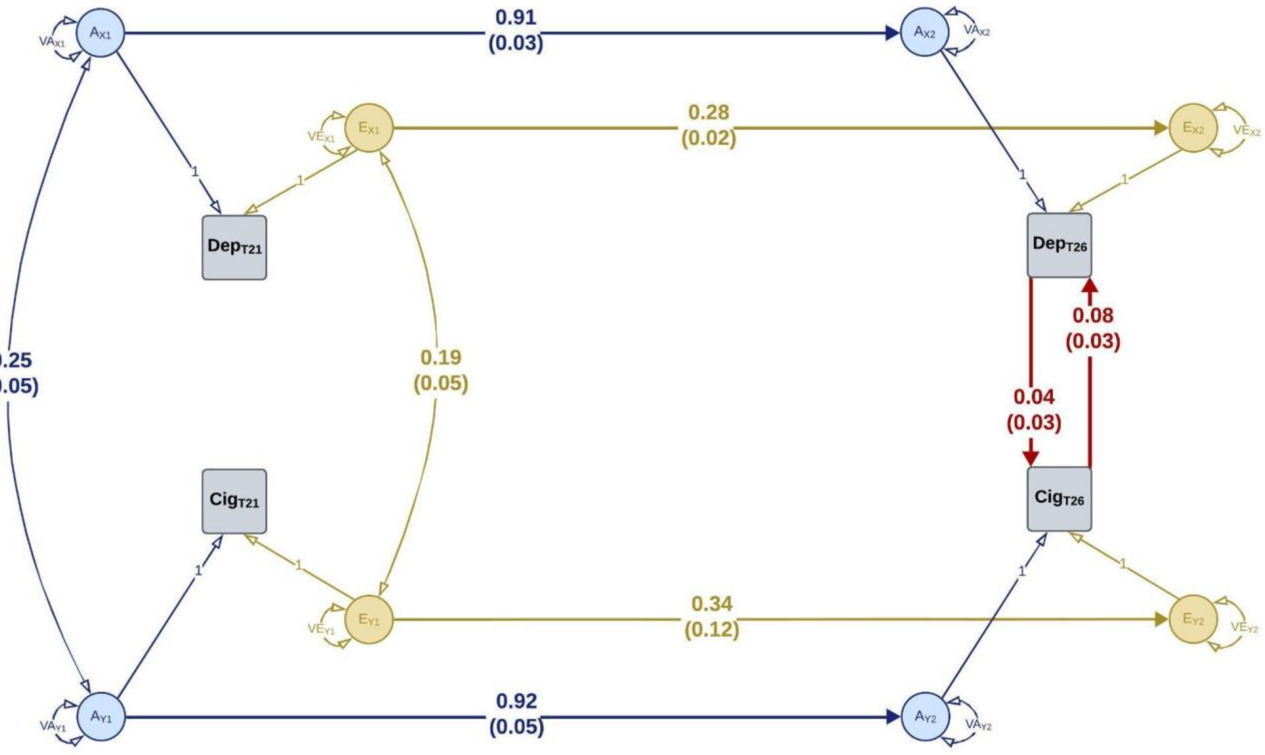

**Figure S4. Biometrical CLPM with Bidirectional Proximal Causation**

*Note. Standardized estimates of selected parameters are shown. The dashed line indicates a non-significant estimate. All parameter estimates are shown in **Supplementary Table S8**.*
